# Digital readiness for cardiovascular care in high-burden US communities with cardiology workforce constraints

**DOI:** 10.64898/2026.09.11.26362838

**Authors:** Sudheesha Perera, Lovedeep Dhingra, Bruno Batinica, Aline Pedroso, Rohan Khera

## Abstract

Digital health may extend cardiovascular care in communities with limited specialist access, but its effectiveness depends on whether communities have the infrastructure, devices, affordability, and capacity needed to use digital services. We linked national data on cardiometabolic burden, cardiology workforce, and digital readiness across 82,999 US census tracts to identify communities where digital cardiovascular care could be deployed and those where enabling investment may be needed first. Among 49,948 tracts in 2,582 counties with no cardiologist or declining per-capita cardiologist supply, 26,517 had above-average cardiometabolic burden. Of these, 4,765 had digital readiness at or above the national median and were classified as deployment-priority tracts, whereas 21,752 had lower readiness and were classified as investment-priority tracts. Investment-priority tracts clustered in the rural Southwest, Deep South, Mississippi Delta, and Gulf and Florida metropolitan areas, whereas deployment-priority tracts were dispersed across urban counties nationwide. Contrasting classifications frequently occurred among neighboring census tracts within the same metropolitan area, highlighting geographic variation not captured by county-level targeting. Findings were broadly consistent across eight additional analyses varying measures of readiness, workforce access, and classification thresholds. We also developed public interactive dashboards that allow users to examine local patterns and modify classification thresholds. These findings provide a national framework for distinguishing communities where digital cardiovascular care may be deployed from those where infrastructure and adoption support may be needed to enable equitable implementation.

## INTRODUCTION

Cardiovascular disease remains the leading cause of death in the United States, yet access to cardiovascular care is geographically uneven.^1^ Nearly half of US counties have no practicing cardiologist, and residents of these counties have a greater burden of cardiovascular risk factors and mortality.^2^ Digital health technologies, including telecardiology, remote monitoring and wearable devices, may extend specialist capacity beyond conventional sites of care.^3–5^ Randomized trials have shown that selected remote-care strategies improve outcomes in heart failure and hypertension.^6–9^ These findings have increased interest in digital health as a strategy for communities with limited specialist access.^3^

Digital care, however, depends on local connectivity, device access, affordability and the skills needed to use digital services.^10,11^ During the COVID-19 pandemic, older adults, people with lower incomes and residents of less connected communities were less likely to complete video visits, including for cardiovascular care.^12,13^ Because high disease burden and low digital readiness often overlap, programs deployed without regard to these conditions may reproduce or widen existing inequities.^11,13^ Yet no national assessment has jointly mapped cardiometabolic burden, cardiology workforce constraints and digital readiness at the community level.

We linked national public datasets to map these dimensions at the census-tract level and identify two complementary priority groups among communities with an absent or declining cardiology workforce: higher-readiness tracts that may be positioned for digital health deployment, and lower-readiness tracts that may require parallel infrastructure and adoption support. We assessed whether these classifications were stable across alternative definitions of readiness, workforce access and thresholds. We also developed interactive dashboards that allow users to explore geographic patterns and modify the classification thresholds.

## RESULTS

The analysis drew on three public datasets: county cardiology workforce counts from the HRSA Area Health Resources Files, tract-level cardiometabolic burden from the CDC PLACES project and tract-level digital readiness from the Purdue Digital Divide Index (DDI). A county counted as workforce-constrained if it had no cardiologist in 2023 or lost cardiologists per capita from 2010 to 2023. Among 82,999 analyzed census tracts in 3,140 counties, 49,948 tracts were located in 2,582 workforce-constrained counties **[Figure 1a–e]**. This included 11,234 tracts in 2,002 counties with no cardiologist in 2023 and 38,714 tracts in 580 additional counties with a declining number of cardiologists per 100,000 residents.

**Figure 1.**
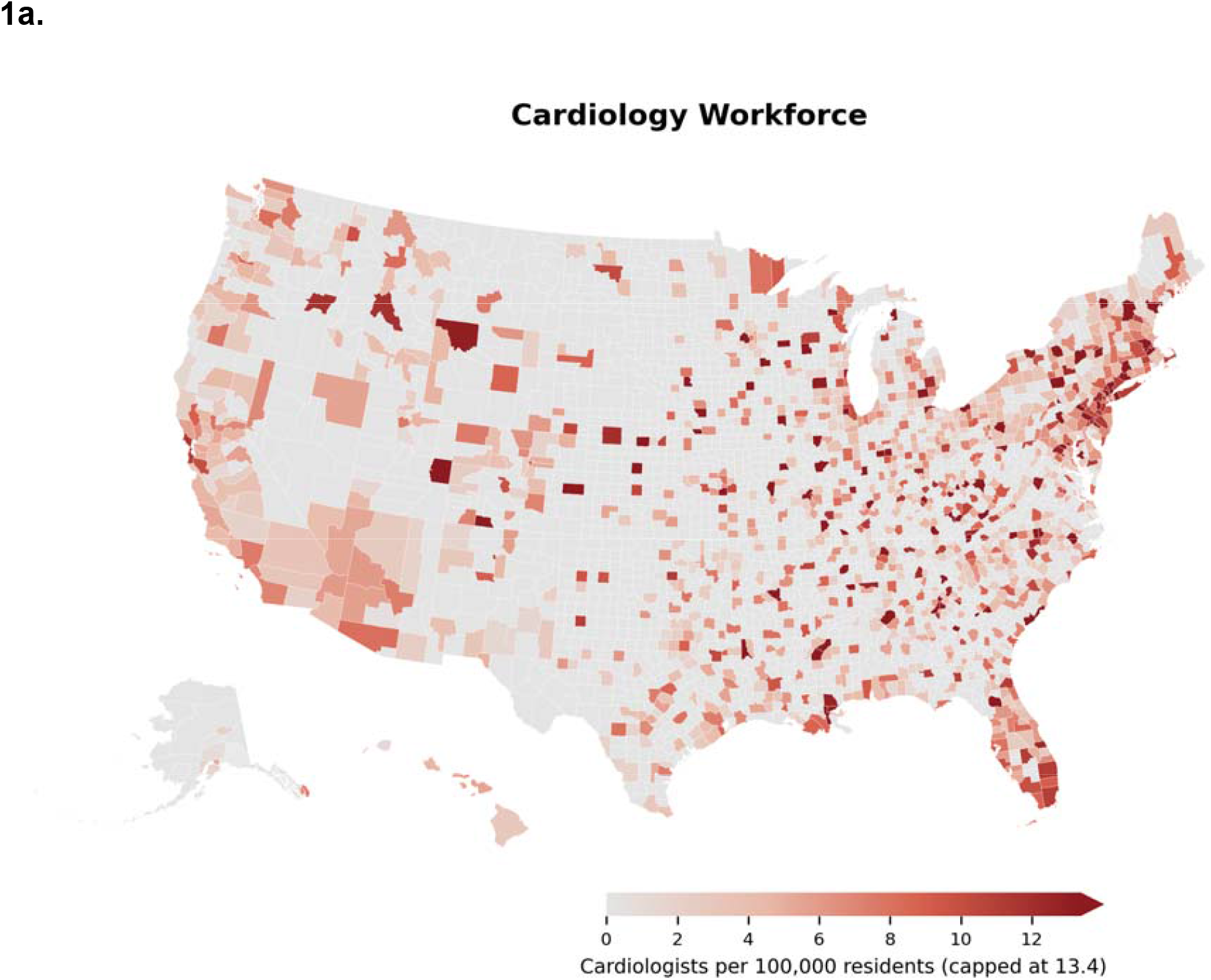
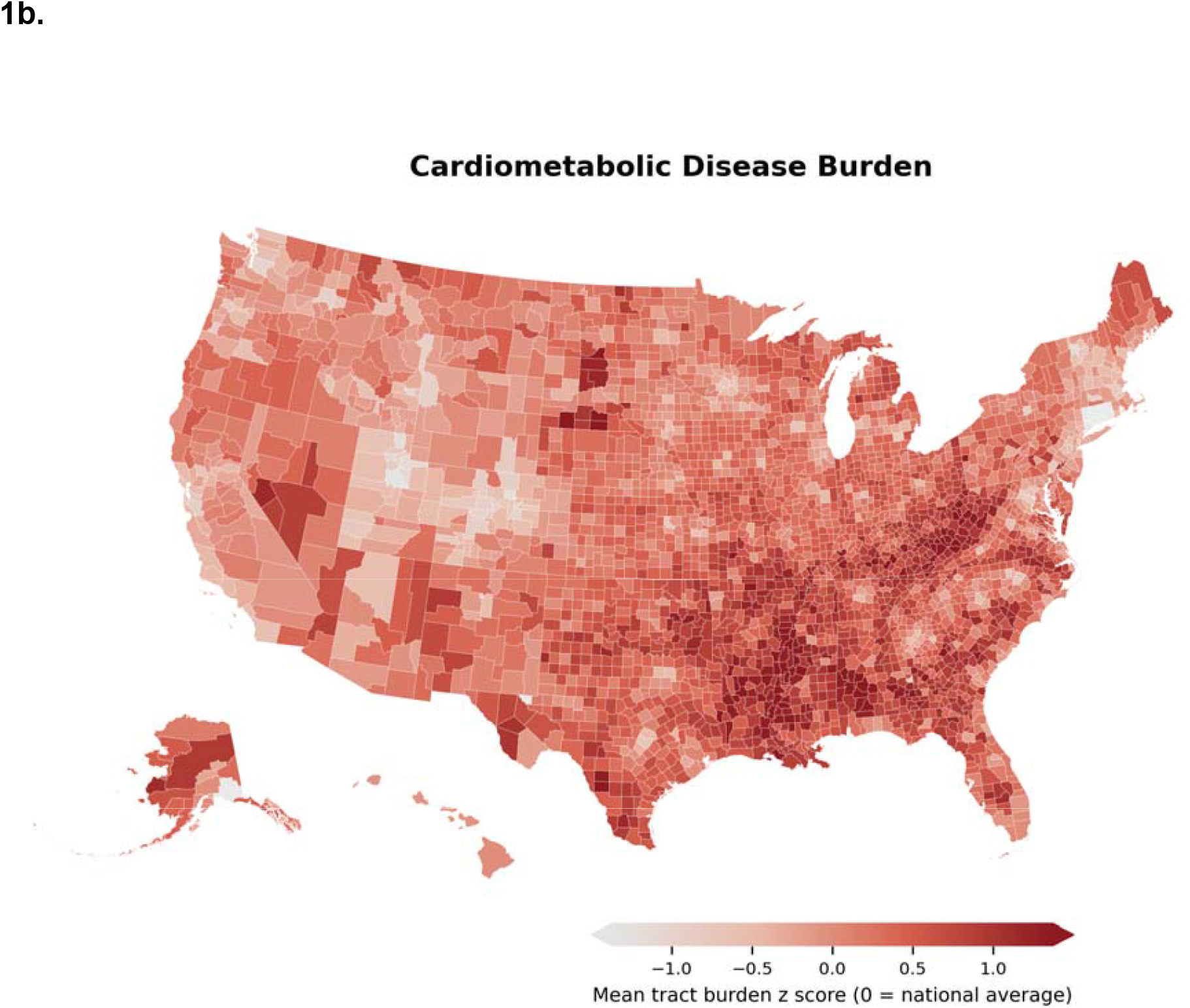
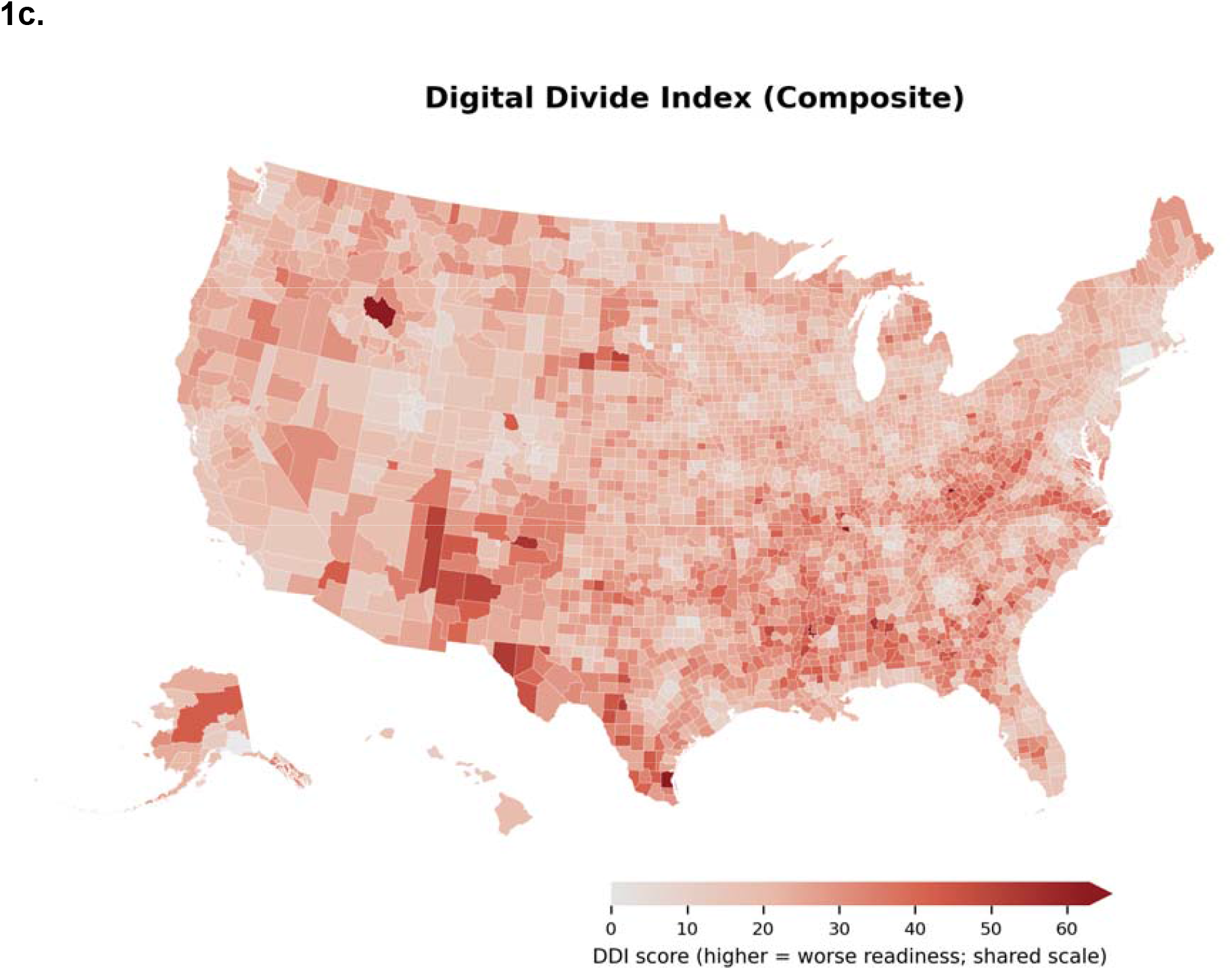
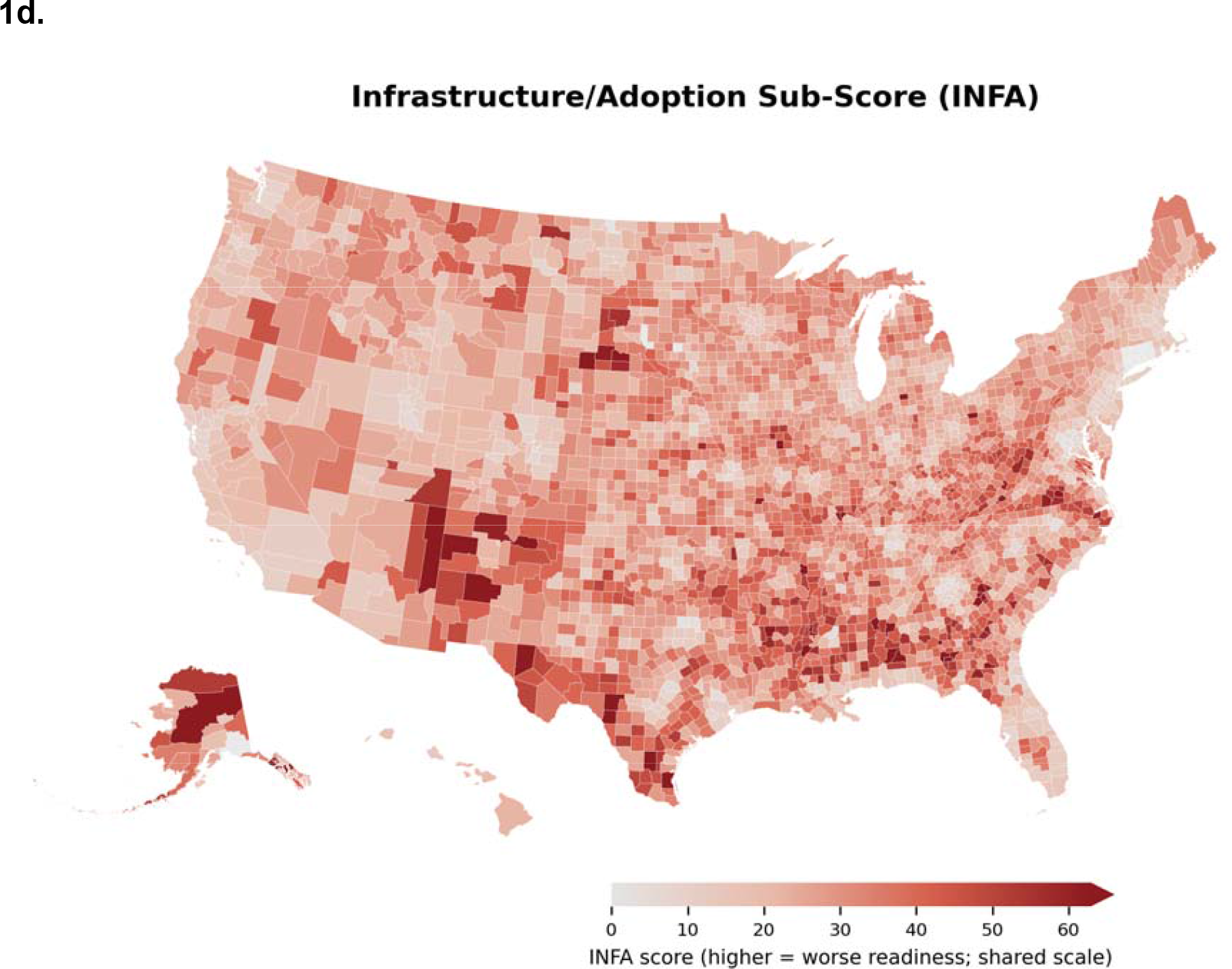
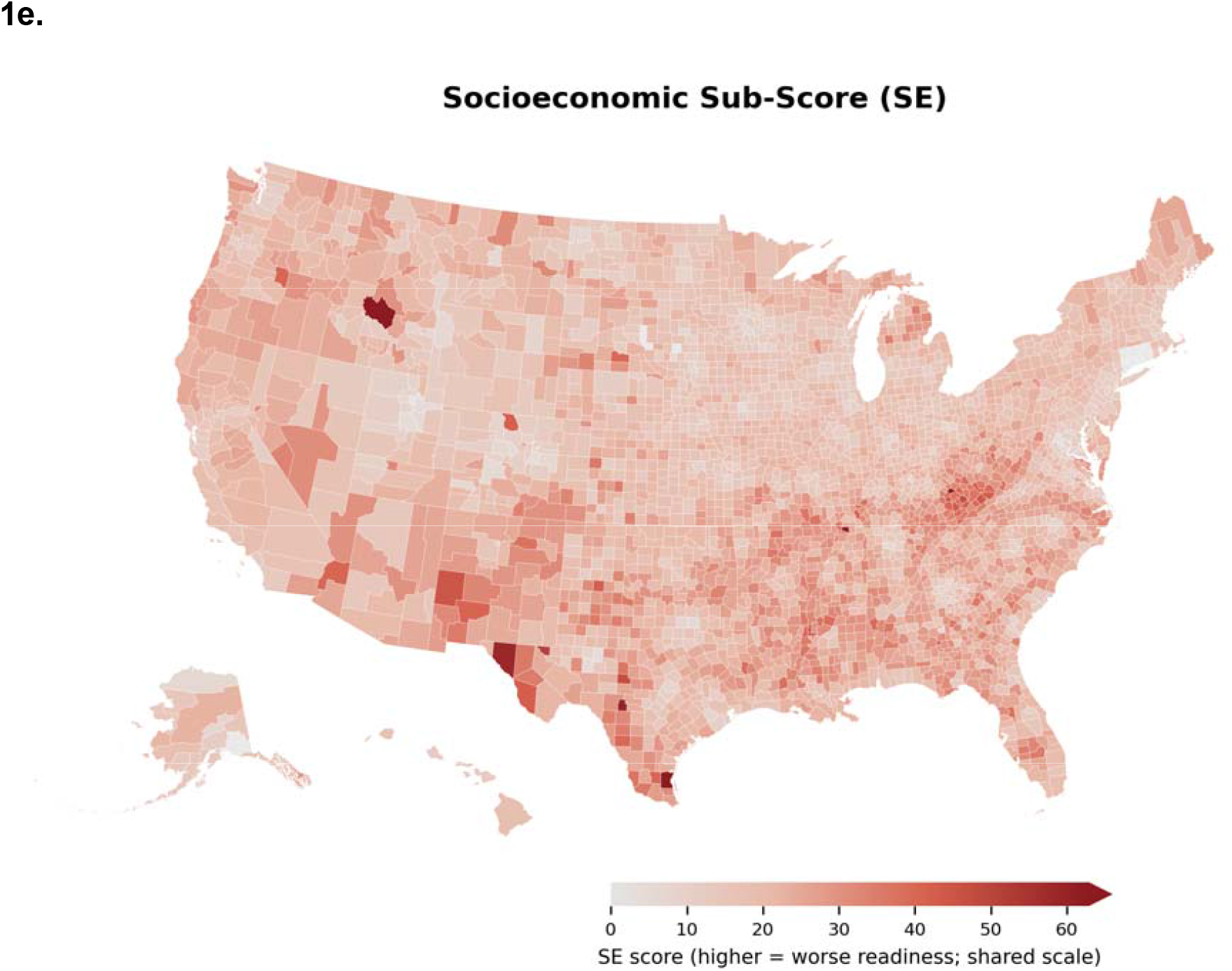
National distributions of cardiology workforce, cardiometabolic burden and digital readiness. (a) Nonfederal cardiologists per 100,000 residents in 2023 (Area Health Resources Files), capped at the 97th percentile (13.4 per 100,000). (b) Mean tract-level cardiometabolic burden z score within each county (PLACES, 2025 release); 0 denotes the national mean. (c) Digital Divide Index (DDI), 2022. (d) DDI infrastructure/adoption subscore, 2022. (e) DDI socioeconomic subscore, 2022. In a, darker red denotes more cardiologists; in b–e, darker red denotes greater burden and lower digital readiness. The scale in a is inverted relative to b–e so that the comparatively few counties with substantial workforce supply stand out against the majority that have few or no cardiologists. Alaska and Hawaii are shown as insets; US territories were excluded.

Of these 49,948 tracts, 26,517 had cardiometabolic burden above the national mean **[Figure 2]**. The DDI scores each tract from 0 to 100 on broadband and device access, internet adoption and socioeconomic barriers. Higher scores mark a larger digital divide and therefore lower readiness. Using the national median DDI (18.8) to distinguish higher from lower readiness, 4,765 tracts were classified as deployment priority and 21,752 as investment priority, representing 19.9 million and 74.1 million residents, respectively **[Table 1]**. Investment-priority tracts had greater burden (median z score 0.55, IQR 0.28–0.90) and lower readiness (median DDI 25.6, IQR 22.3–29.9) than deployment-priority tracts (median z score 0.21, IQR 0.09–0.39; median DDI 16.6, IQR 15.0–17.8), and were more often located in counties without a cardiologist (39.8% versus 14.8% of tracts). The groups also differed in settlement pattern: 18.2% of investment-priority residents lived in noncore rural counties compared with 2.4% of deployment-priority residents, whereas 33.8% of deployment-priority residents lived in large fringe metropolitan counties compared with 14.1% of investment-priority residents. Within the investment-priority group, the median tract had 87.2% of residents living in urban areas (IQR 0.0–100.0), indicating that lower readiness occurred in both rural and urban settings. The 25 highest-ranked tracts in each group are presented in **Tables S1 and S2**. Kentucky and Pennsylvania tracts retained their priority classification but were excluded from ranked lists because burden estimates in those states were based on one measure per the CDC Places data release.

**Figure 2.**
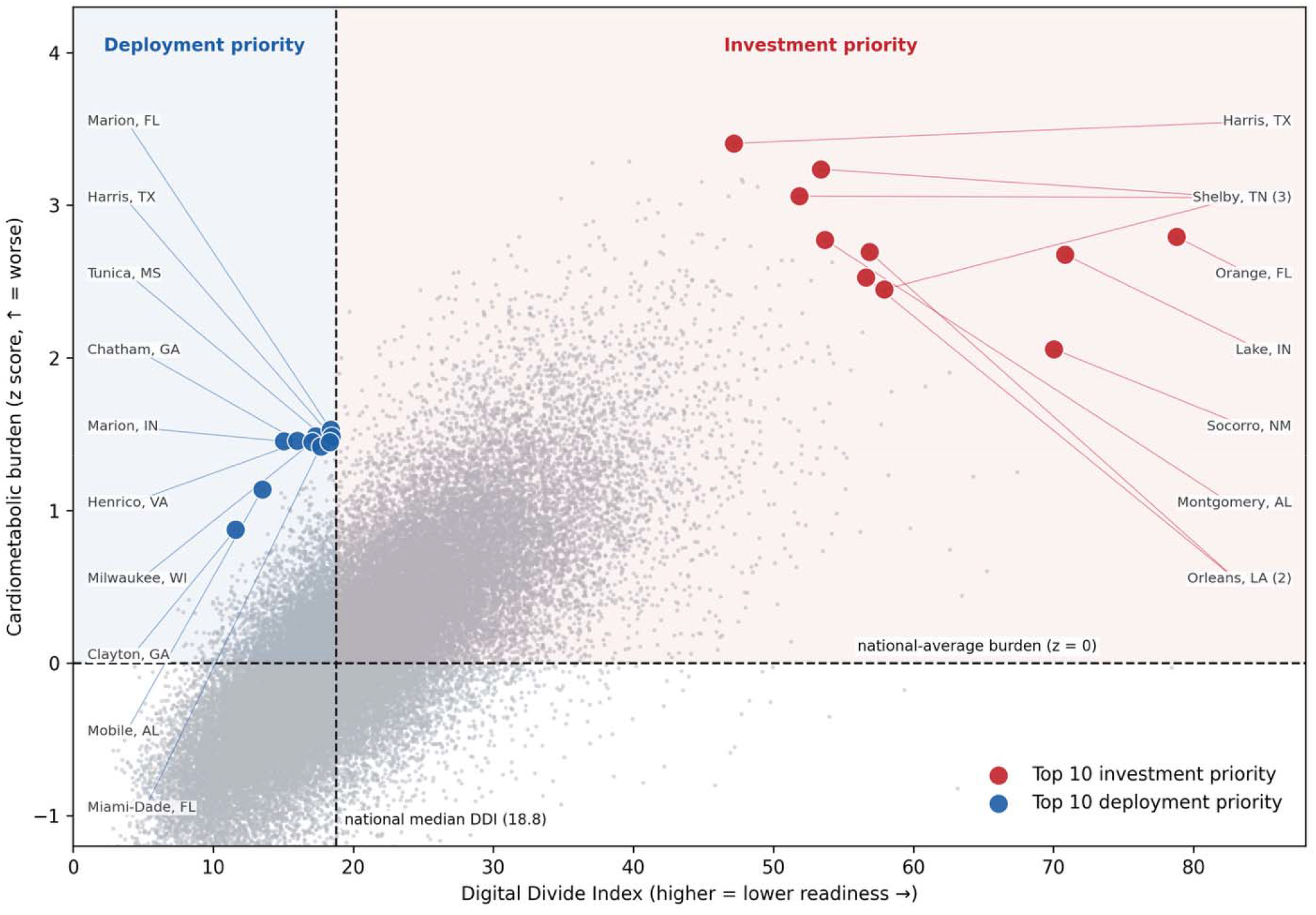
Burden–readiness decision space among workforce-constrained census tracts. Each gray point represents one tract, positioned by DDI (x axis; higher values indicate lower readiness) and cardiometabolic burden z score (y axis). Dashed lines denote the national median DDI (18.8) and national mean burden (z = 0). The upper-left quadrant contains high-burden, higher-readiness deployment-priority tracts; the upper-right contains high-burden, lower-readiness investment-priority tracts. Colored circles identify the 10 highest-ranked tracts in each group; Tables S1 and S2 list the top 25. Kentucky and Pennsylvania tracts were excluded from ranking and from the background distribution because their primary burden estimate was based on one measure; these tracts retain their priority classification and are shown in Fig. 3. Numbers in parentheses after a label give the number of ranked tracts that county contributes.

**Table 1.** Characteristics of census tracts in workforce-constrained counties, by priority group. Tracts are grouped by the 2 × 2 classification of cardiometabolic burden (above versus at or below the national mean) and digital readiness (DDI at or below versus above the national median of 18.8). Residents represented are the sum of tract population from the 2020–2024 American Community Survey 5-year estimates. Values are medians with interquartile ranges across tracts unless otherwise indicated. Urban– rural composition is the percentage of group residents living in counties of each 2023 NCHS urban–rural category. The interquartile range of the urban share spans 0–100% in the investment-priority group because that group is strongly bimodal, comprising both wholly rural and wholly urban tracts. DDI, Digital Divide Index; IQR, interquartile range; NCHS, National Center for Health Statistics.

| Characteristic | Higher burden, higher readiness (deployment priority) | Higher burden, lower readiness (investment priority) | Lower burden, higher readiness | Lower burden, lower readiness | All workforce-constrained |
| --- | --- | --- | --- | --- | --- |
| Tracts, n | 4,765 | 21,752 | 18,116 | 5,315 | 49,948 |
| Residents, n | 19,928,374 | 74,141,508 | 83,476,267 | 20,657,912 | 198,204,061 |
| Median tract population | 3,985 | 3,258 | 4,328 | 3,737 | 3,732 |
| Burden z score, median (IQR) | 0.21 (0.09–0.39) | 0.55 (0.28–0.90) | –0.47 (–0.73 to –0.25) | –0.21 (–0.39 to –0.09) | 0.05 (–0.38 to 0.50) |
| DDI, median (IQR) | 16.6 (15.0–17.8) | 25.6 (22.3–29.9) | 13.6 (11.0–15.9) | 21.3 (19.9–23.6) | 19.6 (14.8–25.0) |
| DDI infrastructure/adoption subscore, median (IQR) | 15.2 (13.1–17.3) | 24.6 (20.7–29.4) | 13.4 (11.3–15.6) | 20.5 (18.0–23.6) | 18.4 (14.0–24.3) |
| DDI socioeconomic subscore, median (IQR) | 10.4 (9.2–11.7) | 14.9 (12.8–17.4) | 8.5 (6.9–10.1) | 13.0 (11.6–14.7) | 11.8 (9.2–14.7) |
| Cardiologists per 100,000, median (IQR) | 4.55 (2.18–8.65) | 2.58 (0.00–6.71) | 6.61 (3.77–9.49) | 5.15 (2.40–8.62) | 4.57 (1.16–8.50) |
| Tracts in counties with no cardiologist, % | 14.8 | 39.8 | 5.4 | 16.9 | 22.5 |
| Burden z score, population-weighted median (IQR) | 0.20 (0.08–0.38) | 0.51 (0.26–0.85) | –0.47 (–0.72 to –0.25) | –0.21 (–0.38 to –0.09) | –0.04 (–0.43 to 0.40) |
| DDI, population-weighted median (IQR) | 16.5 (14.8–17.7) | 25.0 (22.0–29.1) | 13.3 (10.8–15.7) | 21.2 (19.9–23.3) | 18.4 (13.9–23.7) |
| Residents living in urban areas, % of group | 81.6 | 59.4 | 91.8 | 79.2 | 77.4 |
| Urban share of tract population, median % (IQR) | 100.0 (78.6–100.0) | 87.2 (0.0–100.0) | 100.0 (97.7–100.0) | 100.0 (60.8–100.0) | 100.0 (40.0–100.0) |
| **NCHS urban–rural category, % of residents** |  |  |  |  |  |
| Large central metro | 20.0 | 15.5 | 33.2 | 22.7 | 24.2 |
| Large fringe metro | 33.8 | 14.1 | 31.9 | 16.6 | 23.8 |
| Medium metro | 25.9 | 23.9 | 25.5 | 30.5 | 25.4 |
| Small metro | 10.3 | 12.0 | 6.3 | 14.1 | 9.6 |
| Micropolitan | 7.5 | 16.3 | 2.6 | 11.0 | 9.1 |
| Noncore (rural) | 2.4 | 18.2 | 0.5 | 5.1 | 7.8 |

Investment-priority tracts clustered in the rural Southwest, Deep South, Mississippi Delta and Gulf and Florida metropolitan areas **[Figure 3a]**. Deployment-priority tracts were more geographically dispersed and concentrated in urban counties, including Indianapolis, Houston, Milwaukee and metropolitan areas across the South and Mid-Atlantic **[Figure 3b]**. Investment-priority tracts covered a substantially larger land area, reflecting their rural distribution, whereas the two groups differed less in population than in area **[Figure 3c]**.

**Figure 3.**
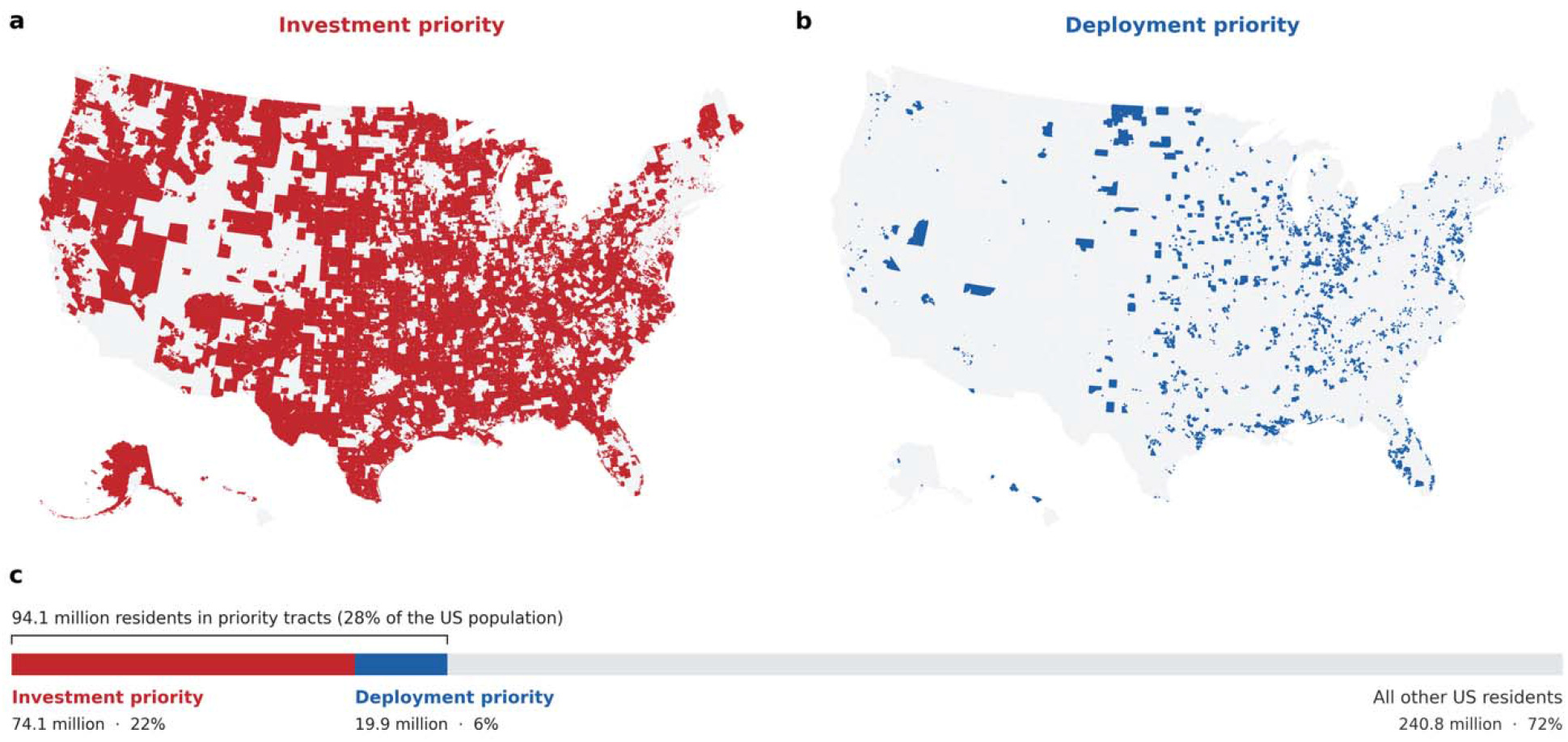
National distribution of priority tracts and the population they represent. (a) Census tracts classified as investment priority (red) and (b) deployment priority (blue). The colored regions mark the locations of census tracts and do not reflect the size of their populations: investment-priority tracts occupy 57% of the land area of the contiguous United States compared with 3.8% for deployment-priority tracts, because they are predominantly rural. (c) Residents of priority tracts as a share of the US population, with 74.1 million (22%) in investment-priority tracts and 19.9 million (6%) in deployment-priority tracts, together 28% of the population. Kentucky and Pennsylvania tracts are shown because they retained their priority classification, although they were excluded from the ranked lists. Alaska and Hawaii are shown as insets.

### Metropolitan Area Patterns

Within metropolitan areas, deployment-and investment-priority tracts often occurred in adjacent neighborhoods **[Figure 4a–e]**. Delaware County, Pennsylvania, contained a large contiguous cluster of deployment-priority tracts; these tracts were excluded from national rankings because the primary Pennsylvania burden estimate was based on one measure, although 16 of 19 remained above the national mean when burden was reconstructed from the most recent full-measure PLACES release **[Table S3]**. The central counties of the Philadelphia, Dallas, Washington and Boston metropolitan areas were outside the workforce-constrained pool, whereas Cook County, Illinois, was included and contained both priority groups.

**Figure 4.**
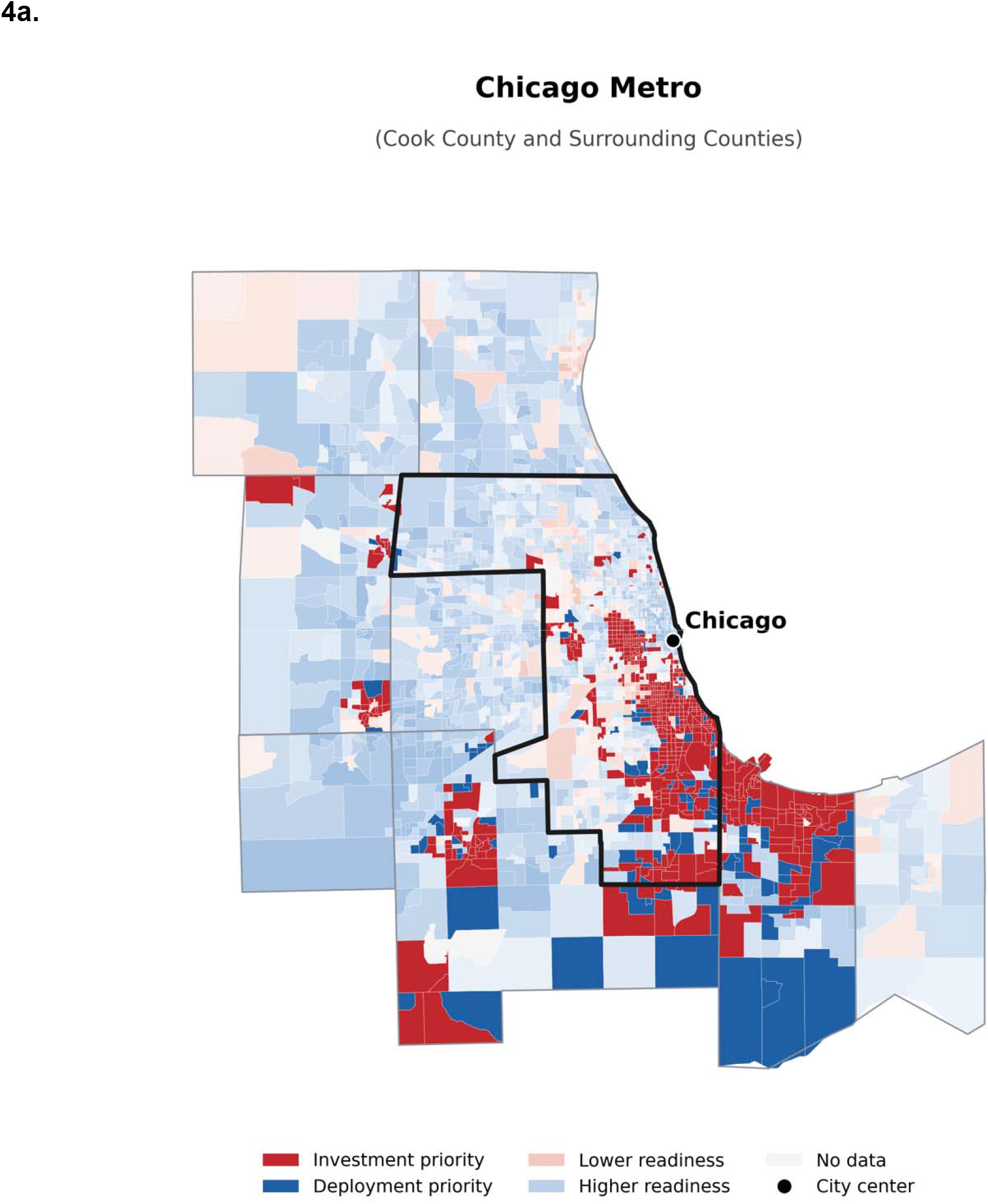
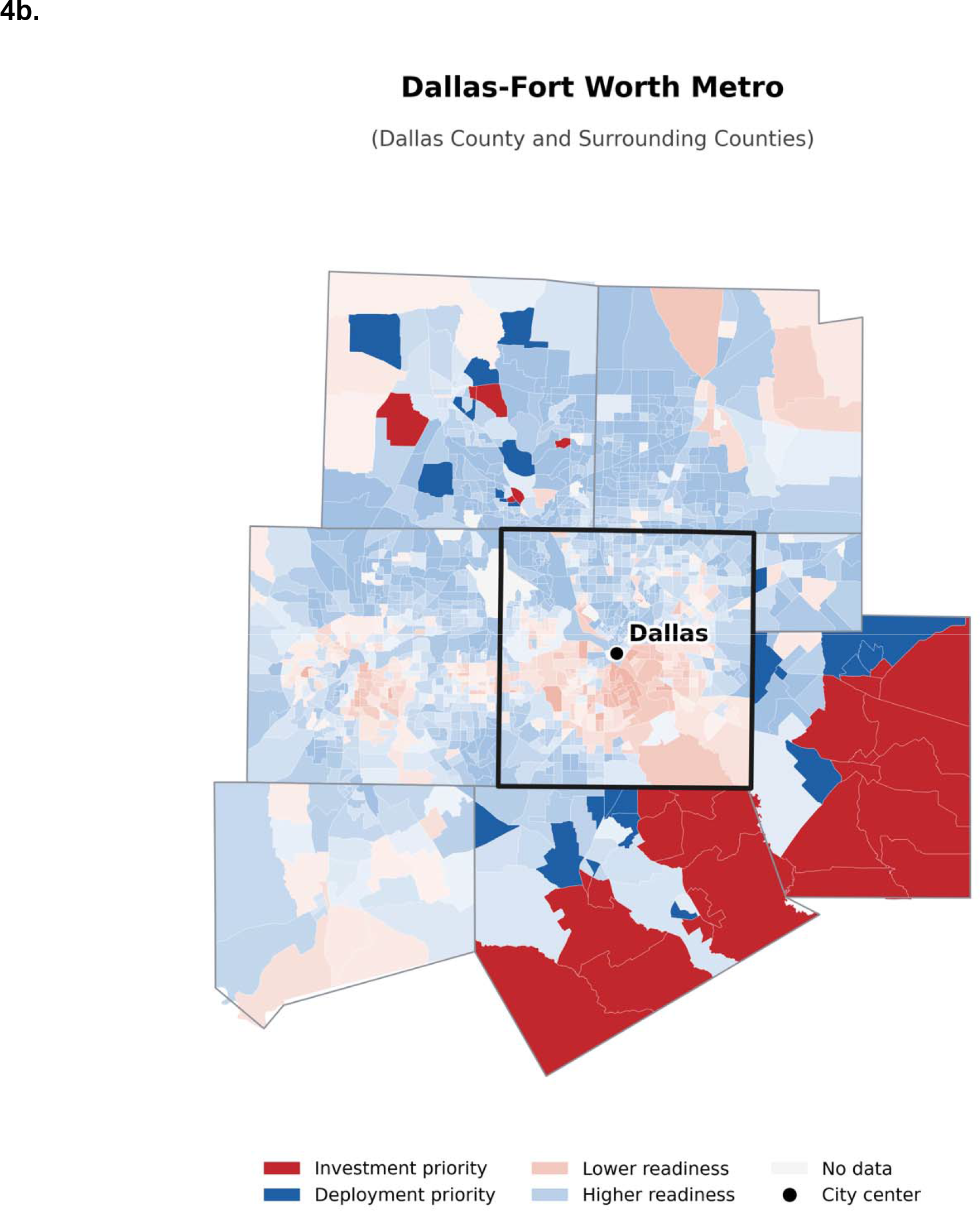
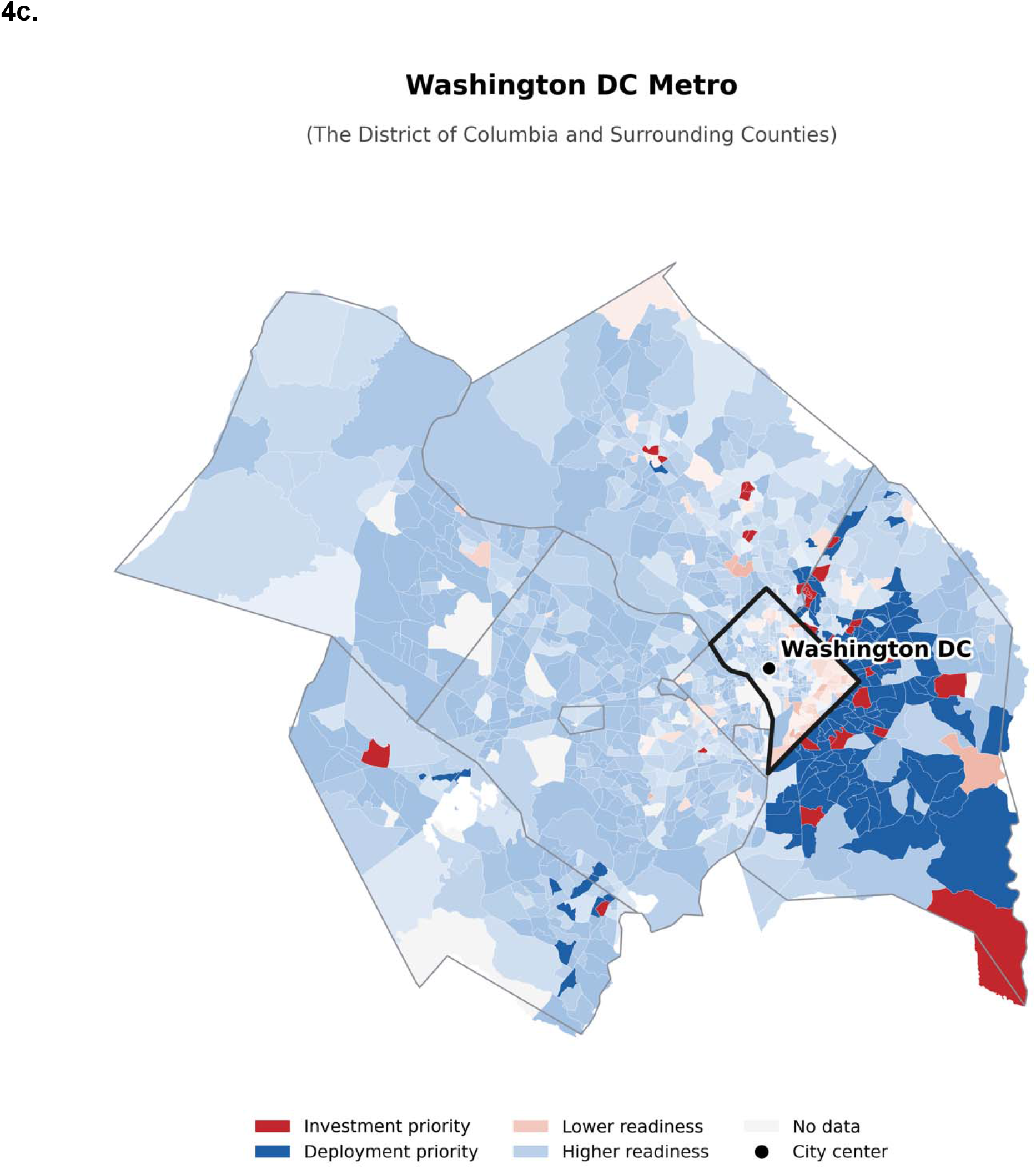
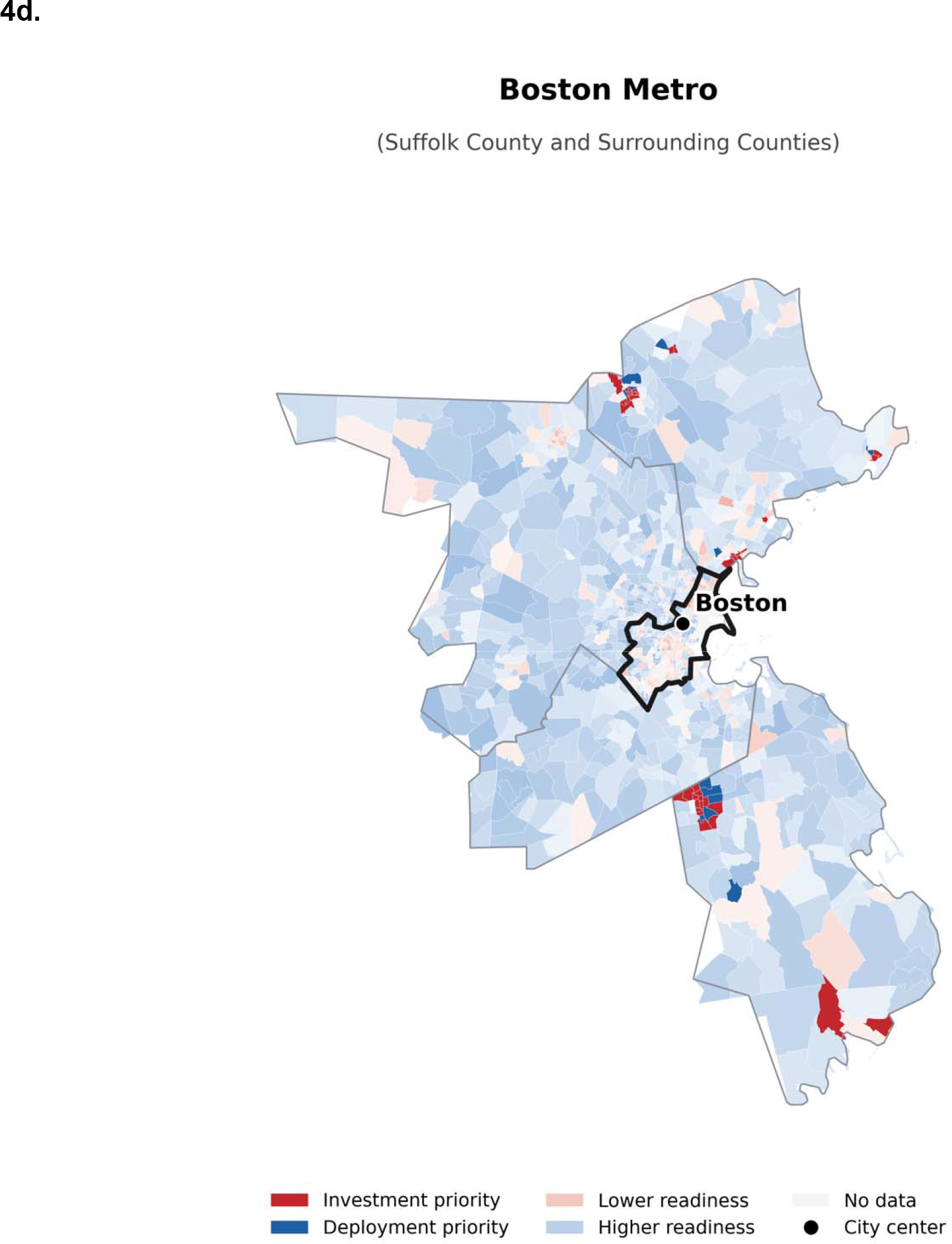
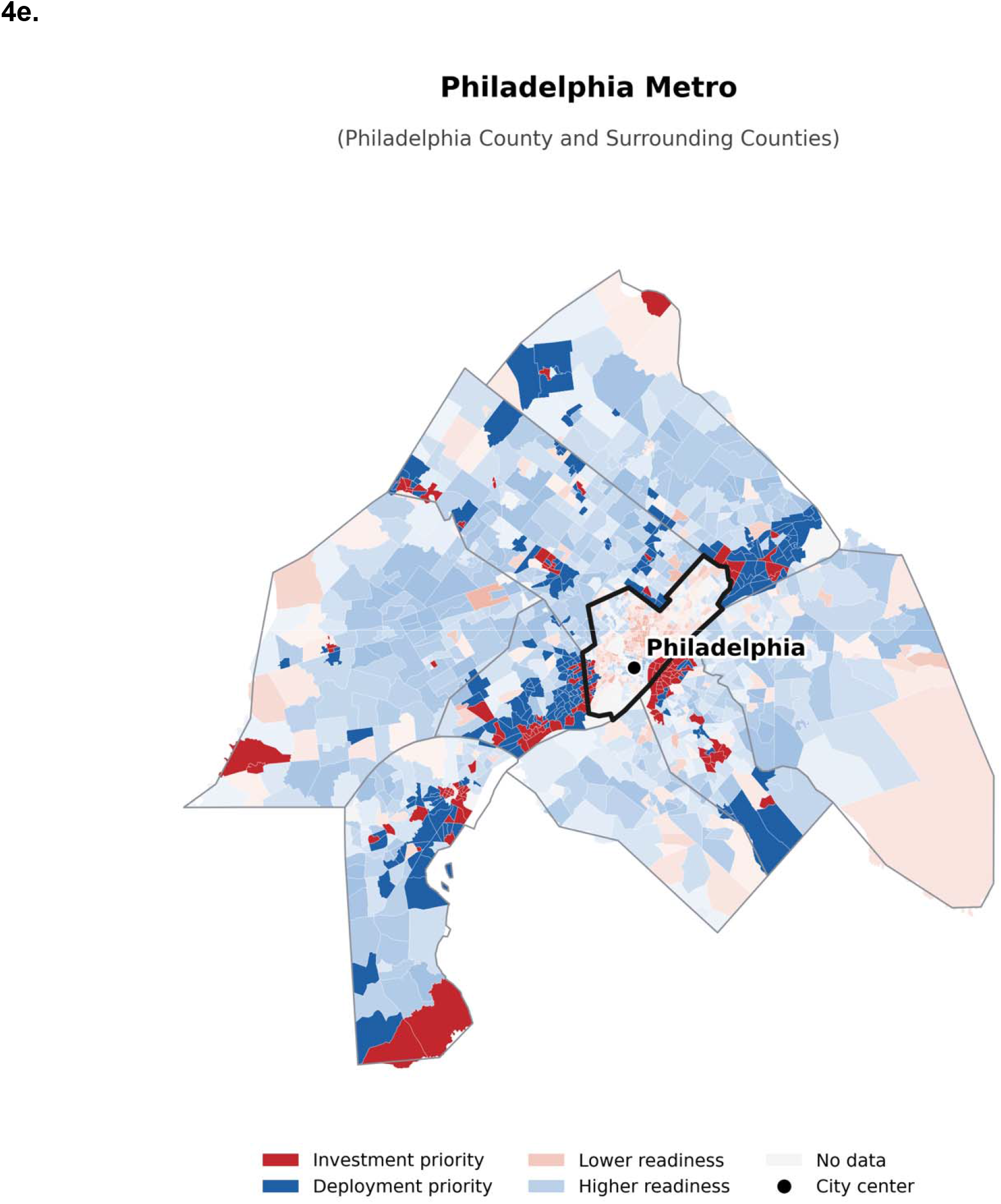
Neighborhood-level variation across five metropolitan areas. Census tracts are colored by the primary classification: red denotes investment priority and blue deployment priority. Light red denotes lower readiness and light blue higher readiness among tracts outside the two high-burden priority groups; tracts without data are gray. Black outlines identify the central county and black circles the city center. (a) Chicago (Cook County, Illinois); (b) Dallas–Fort Worth (Dallas County, Texas); (c) Washington, DC (District of Columbia); (d) Boston (Suffolk County, Massachusetts); and (e) Philadelphia (Philadelphia County, Pennsylvania). The central counties of the Dallas, Washington, Boston and Philadelphia metropolitan areas were outside the workforce-constrained pool; priority tracts were located in surrounding counties.

### Sensitivity Analyses

Alternative readiness definitions yielded high overall agreement with the primary classification but greater variation in the highest-ranked deployment-priority tracts. A six-indicator readiness index correlated with the DDI (Pearson r = 0.89) and produced 93.2% box agreement; adding FCC broadband availability increased agreement to 93.6% **[Tables S4 and S5]**. The infrastructure/adoption and socioeconomic subscores produced 94.0% and 92.2% agreement, respectively, but emphasized different geographies **[Tables S6–S9]**. Removing the workforce screen expanded the classification to 31,975 investment-and 7,560 deployment-priority tracts and shifted the highest-ranked tracts toward urban counties excluded by the primary screen **[Tables S10 and S11]**. All 10 leading investment-priority tracts remained classified under every stricter burden/readiness threshold combination, whereas the leading deployment-priority tracts were sensitive to readiness thresholds below the median **[Table S12]**. Allowing cross-county workforce access produced 92.7% box agreement and retained 9 of the top 10 tracts in each group **[Tables S13 and S14]**. Adding cardiology-affiliated nurse practitioners and physician assistants produced 97.1% agreement, with all 10 leading investment-priority tracts and all 10 leading deployment-priority tracts retained **[Tables S15 and S16]**. Rebuilding the pool around the primary care workforce retained 51.3% of investment-priority tracts and shifted leading deployment-priority tracts **[Tables S17 and S18]**. In Kentucky and Pennsylvania, the reconstructed 10-measure burden correlated with the primary burden (Pearson r = 0.91), and 16 of 19 leading Delaware County tracts remained above the national mean **[Table S3]**.

### Interactive Dashboards

The dashboards provide tract-level views of all 82,999 analyzed tracts. The national map extends the static maps to every county and census tract, and the implementation decision space reproduces the burden–readiness matrix interactively, allowing users to modify thresholds, compare geographies and identify leading tracts. Both dashboards are publicly available at https://digital-readiness.cards-lab.org/ (national map: https://digital-readiness.cards-lab.org/map; implementation decision space, https://digital-readiness.cards-lab.org/decision-space) and were last updated on August 18, 2026. Both dashboards were designed to meet Web Content Accessibility Guidelines 2.1 Level AA, including keyboard navigation, text alternatives for map and plot elements, and color palettes selected to remain legible under common forms of color vision deficiency.

## DISCUSSION

This national census-tract analysis identified two distinct priority profiles among high-burden communities with an absent or declining cardiology workforce. Higher-readiness deployment-priority tracts were dispersed across urban counties, whereas lower-readiness investment-priority tracts clustered in the rural Southwest, Deep South, Mississippi Delta and Gulf and Florida metropolitan areas. Broad classifications were stable across alternative readiness and workforce definitions, although the highest-ranked deployment-priority tracts varied with the readiness measure. Two interactive dashboards demonstrate the ability to operationalize this framework at the tract level and allow users to examine how local priorities change under alternative thresholds.

The geography underscores that limited specialist supply and digital readiness are not synonymous. Investment-priority tracts clustered in the rural Southwest, Deep South and Mississippi Delta, including tribal communities where cardiovascular risk and infrastructure deficits are well documented.^14,15^ Deployment-priority tracts were more often urban, but higher tract-level readiness did not imply universal access: cardiovascular telemedicine studies during the COVID-19 pandemic found lower video-visit completion among older adults, people with lower incomes and patients with a preferred language other than English.^12,13^ Tract-level readiness should therefore be interpreted as a starting point for local implementation assessment rather than evidence that all residents can use a given technology.

Variation across readiness measures also reflects the multidimensional nature of digital access. A community may have adequate broadband but face barriers related to devices, affordability, digital skills or language access, or the reverse. The DDI composite balances infrastructure and socioeconomic dimensions, whereas its subscores may help identify whether implementation should prioritize broadband and device access or affordability and digital navigation. Intervention design should therefore be matched to the specific local barrier rather than to a binary designation of readiness.

The framework separates policy levers that are often treated as interchangeable. Lower-readiness communities may require broadband build-out, affordability support, device access and digital navigation before or alongside clinical deployment; higher-readiness communities may be more constrained by reimbursement, workflow integration and implementation capacity. Several key federal programs address different parts of this pathway. The $42.45 billion Broadband Equity, Access, and Deployment (BEAD) program funds infrastructure investment.^16^ Meanwhile, the FCC Rural Health Care Program, capped at $744 million for funding year 2026, subsidizes provider connectivity.^17,18^ The Connected Care Pilot Program, which covered 85% of connectivity costs for 111 selected projects, concluded in 2025.^19^ Separately, the Medicare telehealth flexibilities, extended through 2027, together with state Medicaid policies, govern reimbursement.^20,21^ The Rural Health Transformation Program, which distributed $50 billion to all 50 states for 2026–2030, explicitly supports telehealth, remote patient monitoring and digital tools, with states determining which rural communities receive them.^22^ Recently, affordability support has contracted: the Affordable Connectivity Program, which reached 23 million households, 72% of whom reported using it to schedule or attend healthcare appointments, ended in June 2024, leaving Lifeline’s $9.25 monthly benefit as the principal subsidy.^23–25^ Because program funding and eligibility continue to change, with roughly $21 billion in BEAD savings unallocated and Digital Equity Act programs partially restored under court order, geographic targeting should be paired with current policy conditions rather than treated as a static designation.^26–28^

Some limitations should be considered. First, source data lagged the present, with workforce measures extending through 2023, the Divide Index through 2022, and burden estimates through the 2023 Behavioral Risk Factor Surveillance System, with short sleep duration through 2022, as published in the 2025 CDC PLACES census tract release. These conditions move slowly: county cardiologist counts were overall nearly identical between consecutive years (Pearson r = 0.999) and 98.6% of counties retained the same no-cardiologist status between 2022 and 2023. The three data sources are closely aligned in time and describe the post-pandemic period. Second, workforce was measured at the county level and assigned to census tracts; physician counts also do not capture appointment availability, subspecialty mix, cross-county travel or existing telecardiology capacity. The 25-mile analysis addressed but did not eliminate this limitation. Third, public data were incomplete for Connecticut, Kentucky and Pennsylvania, although analyses using alternative geographies and the most recent full-measure PLACES release produced broadly consistent classifications. Fourth, the DDI measures general digital disadvantage rather than uptake or effectiveness of a specific cardiovascular intervention, and tract-level conditions do not establish individual access or skills. Finally, the relative thresholds and priority labels were not validated against implementation outcomes. The dashboards should therefore support, rather than replace, local assessment.

US communities with high cardiometabolic burden and cardiology workforce constraints differ substantially in digital readiness. A tract-level prioritization framework and public dashboards distinguish areas where digital cardiovascular programs may be more feasible from those where infrastructure and adoption support should accompany deployment.

## ONLINE METHODS

### Data Sources and Study Population

We linked three national, publicly available datasets at the census-tract level. County-level cardiology workforce counts came from the Health Resources and Services Administration Area Health Resources Files (2010–2023); tract-level cardiometabolic burden came from the Centers for Disease Control and Prevention PLACES project (2025 census tract release, which reports 2023 Behavioral Risk Factor Surveillance System data, with short sleep duration carried forward from 2022); and digital readiness came from the Purdue Digital Divide Index, which is derived from American Community Survey and Federal Communications Commission data. We included US census tracts with complete data on cardiometabolic burden and digital readiness and assigned county-level workforce measures to each tract. Because the analysis used only public, deidentified data, institutional review board oversight was not required.

### Cardiology Workforce and Cardiometabolic Burden

We measured the cardiology workforce using county-level counts of nonfederal cardiovascular disease physicians. We characterized each county by whether any cardiologist was present in 2023 and by the 2010–2023 trend in cardiologists per 100,000 residents, using every year of county cardiology counts available across the AHRF releases. HRSA publishes this variable for only two years per release. We did not impute values for years not covered by these releases. In counties averaging at least three cardiologists per year with at least three observed years, decline was defined by a negative ordinary least squares slope of the per-capita rate regressed on calendar year. In counties averaging fewer than three, where a fitted slope is unstable, decline was defined instead as a lower per-capita rate in the last observed year than in the first. These criteria were specified when the workforce panel was constructed and applied unchanged in all subsequent analyses. We measured cardiometabolic burden using a tract-level composite z score across 10 conditions and risk factors: hypertension, high cholesterol, coronary heart disease, stroke, diabetes, obesity, current smoking, physical inactivity, short sleep duration and binge drinking. For each measure, we used the age-adjusted prevalence reported in the 2025 PLACES census tract release, standardized values across tracts and averaged the resulting z scores.

### Digital Readiness

Digital readiness was measured with the Purdue Digital Divide Index (DDI), a tract-level composite of infrastructure, adoption and socioeconomic barriers.^29^ The DDI ranges from 0 to 100, with higher values indicating a larger digital divide and therefore lower readiness. The infrastructure and adoption subscore includes fixed broadband availability, household computer access, home internet subscription and advertised download and upload speeds. The socioeconomic subscore includes the proportion of residents aged 65 years or older, the proportion with less than a high-school education, poverty, disability and the Internet Income Ratio. Inputs are derived from the American Community Survey and Federal Communications Commission.^29^

We selected the DDI because it combines infrastructure and socioeconomic dimensions at census-tract resolution using transparent, public inputs.^29^ Prior studies have associated the index with telehealth use and video-visit completion.^30–32^ Deprivation and vulnerability indices, including the Area Deprivation Index and Social Vulnerability Index, do not incorporate broadband or device measures, whereas broadband availability alone does not capture affordability, device access or other barriers to adoption.^33,34^

### Priority Group Definitions

We first defined a workforce-constrained pool comprising counties with no cardiologist in 2023 or a declining number of cardiologists per 100,000 residents over 2010–2023. Within this pool, tracts were classified on two axes using national benchmarks. High burden was defined as a composite burden z score above the national mean, and higher readiness as a DDI at or below the national median. The resulting 2 × 2 framework designated high-burden, higher-readiness tracts as deployment priority and high-burden, lower-readiness tracts as investment priority. These labels indicate relative priorities under the chosen thresholds and do not establish readiness for any specific intervention. To rank tracts within each group, we standardized burden and DDI within the pool and calculated burden minus DDI for deployment priority and burden plus DDI for investment priority. Kentucky and Pennsylvania reported only 1 of the 10 burden measures; their tracts were retained in the classification but excluded from ranked national lists because their burden estimates were not directly comparable.

### Sensitivity Analyses

We conducted eight analyses to assess whether the prioritization framework depended on the readiness measure, workforce definition, geographic access assumption or classification threshold. First, we reconstructed readiness from six standardized American Community Survey indicators of internet access, computer access, poverty, disability, older age and educational attainment, with and without a seventh measure of terrestrial broadband availability from the FCC Broadband Data Collection. Second, we repeated the classification using the DDI infrastructure/adoption and socioeconomic subscores separately. Third, we applied the framework to all US tracts without the workforce screen. Fourth, we crossed three burden thresholds (above the national mean, top quartile and top decile) with three readiness thresholds (median, quartile and decile). Fifth, we credited counties with cardiologists in counties whose geographic centroids were within 25 miles. Sixth, we expanded the workforce to include cardiology-affiliated nurse practitioners and physician assistants identified in the CMS Doctors and Clinicians file. A practice site was defined as one group at one address, and a nurse practitioner or physician assistant was considered cardiology affiliated when cardiologists comprised at least half of physicians at that site. Seventh, we rebuilt the pool using the primary care physician workforce. Eighth, for Kentucky and Pennsylvania, we reconstructed burden from the 2023 PLACES census tract release, the most recent release in which those states reported all 10 measures, and crosswalked estimates to current tracts.

### Metropolitan Area Analyses

To illustrate within-metropolitan heterogeneity, we mapped tract-level classifications in five metropolitan areas: Chicago, Dallas–Fort Worth, Washington, DC, Boston and Philadelphia. For each, we displayed the county containing the city center and surrounding counties, with each tract shown by priority group and readiness relative to the national median.

### Interactive Dashboards

We developed two public dashboards to support exploration beyond the static figures. The national map displays cardiology workforce, cardiometabolic burden, digital readiness and priority classification for each county and census tract. The implementation decision space plots workforce-constrained tracts by burden and readiness and allows users to modify thresholds and identify leading tracts within a selected geography. Both dashboards use only the public data sources described above. The code to generate and deploy the dashboards is also publicly shared.

### Statistical Analysis

We summarized classifications using counts and geographic distributions. We compared alternative readiness indices using Pearson correlation coefficients. Stability was assessed using box agreement (the proportion of tracts retaining the same classification), retention (the proportion of each primary priority group retaining its label) and top-10 overlap. All thresholds were anchored to national distributions among analyzed tracts. Analyses were performed in Python 3.11. Group-level characteristics were summarized as medians with interquartile ranges across tracts, and the number of residents represented was calculated by summing tract population from the 2020–2024 American Community Survey 5-year estimates. Urban–rural composition was described using the 2023 NCHS Urban–Rural Classification Scheme for Counties and the proportion of tract residents living in urban areas in the 2020 Census. Because classification thresholds were anchored to tract-level national distributions, we also calculated population-weighted medians.

### Data Availability

This study used publicly available data. Cardiology workforce counts were obtained from the HRSA Area Health Resources Files (source: https://data.hrsa.gov/data/download). Cardiometabolic burden estimates were obtained from the CDC PLACES project census tract release for 2025 (source: https://data.cdc.gov/500-Cities-Places/PLACES-Local-Data-for-Better-Health-Census-Tract-D/cwsq-ngmh). The Kentucky and Pennsylvania burden reconstruction used the CDC PLACES 2023 census tract release (source: https://data.cdc.gov/resource/hky2-3tpn.csv), the most recent release in which those states reported all 10 measures. Digital readiness scores were obtained from the Purdue Center for Regional Development Digital Divide Index, 2022–2024 release (available at no cost on request; see https://pcrd.purdue.edu/ddi, accessed 13 July 2026). The primary analysis uses the 2022 census tract vintage. American Community Survey 5-year estimates were retrieved through the Census Bureau application programming interface (https://api.census.gov/data). Broadband availability was derived from the Federal Communications Commission Broadband Data Collection served and unserved location files, vintage 31 December 2024 (https://broadbandmap.fcc.gov/data-download). Cardiology nurse practitioner and physician assistant affiliations were derived from the CMS Doctors and Clinicians National Downloadable File (https://data.cms.gov/provider-data). Urban–rural classification used the NCHS Urban– Rural Classification Scheme for Counties (https://www.cdc.gov/nchs/data-analysis-tools/urban-rural.html) and the 2020 Census Demographic and Housing Characteristics File (https://api.census.gov/data/2020/dec/dhc). Census tract boundaries were obtained from Census Bureau cartographic boundary files.

### Code Availability

All analysis code is publicly available at: https://github.com/CarDS-Yale/digital-readiness-for-cardiovascular-care. The release used for this analysis is archived at https://doi.org/10.5281/zenodo.22702589. The repository contains the complete pipeline from the public source files through the priority classification, sensitivity analyses, tables and figures, together with the source code for both dashboards.

## Data Availability

All data produced are available online. Please refer to the "Data Availability" section of the manuscript.

## Acknowledgements

We thank the Purdue Center for Regional Development and Roberto Gallardo, PhD, for providing the Digital Divide Index data used in this analysis.

## Author Contributions

S.P. conceived the study. S.P. performed the analysis and drafted the manuscript. L.D., B.B. and A.P. contributed to interpretation and critical revision. R.K. supervised the work. All authors approved the final manuscript.

## SUPPLEMENTS

**Table S1.** Twenty-five highest-ranked investment-priority census tracts. Tracts are ranked by the investment-priority score, calculated as standardized burden plus standardized DDI. Tract FIPS is the 11-digit Federal Information Processing Standards code. DDI, Digital Divide Index; INFA, infrastructure/adoption subscore; SE, socioeconomic subscore. Cardiologists per 100,000 residents are measured for the parent county in 2023. DDI scores are from 2022.

| Rank | County | State | Tract FIPS | DDI composite | INFA | SE | Cardiologists per 100,000 | Burden z score |
| --- | --- | --- | --- | --- | --- | --- | --- | --- |
| 1 | Orange | FL | 12095010500 | 78.80 | 71.20 | 43.10 | 8.50 | 2.79 |
| 2 | Lake | IN | 18089011700 | 70.80 | 69.20 | 36.00 | 5.59 | 2.68 |
| 3 | Socorro | NM | 35053940000 | 70.00 | 81.70 | 27.90 | 0.00 | 2.06 |
| 4 | Shelby | TN | 47157005900 | 53.40 | 52.00 | 27.90 | 8.68 | 3.24 |
| 5 | Orleans | LA | 22071000620 | 56.80 | 63.00 | 25.00 | 18.40 | 2.69 |
| 6 | Shelby | TN | 47157005800 | 51.90 | 55.30 | 24.30 | 8.68 | 3.06 |
| 7 | Harris | TX | 48201331400 | 47.20 | 35.90 | 30.80 | 7.36 | 3.41 |
| 8 | Shelby | TN | 47157000600 | 57.90 | 58.70 | 28.70 | 8.68 | 2.45 |
| 9 | Orleans | LA | 22071001751 | 56.60 | 51.60 | 31.40 | 18.40 | 2.53 |
| 10 | Montgomery | AL | 01101002400 | 53.70 | 49.60 | 29.50 | 5.33 | 2.77 |
| 11 | Greene | AL | 01063060000 | 48.90 | 50.30 | 24.10 | 0.00 | 3.12 |
| 12 | Navajo | AZ | 04017942400 | 62.60 | 83.30 | 19.10 | 1.83 | 1.90 |
| 13 | Shelby | TN | 47157005500 | 54.20 | 59.50 | 24.30 | 8.68 | 2.63 |
| 14 | Webster | LA | 22119031900 | 47.70 | 50.40 | 22.80 | 0.00 | 3.17 |
| 15 | Ouachita | LA | 22073010700 | 47.80 | 52.70 | 21.60 | 5.08 | 3.15 |
| 16 | Apache | AZ | 04001942600 | 67.40 | 100.00 | 14.40 | 0.00 | 1.25 |
| 17 | Lake | IN | 18089011900 | 46.80 | 51.80 | 21.10 | 5.59 | 3.06 |
| 18 | Sandoval | NM | 35043940900 | 60.90 | 73.90 | 22.80 | 0.00 | 1.81 |
| 19 | Pulaski | AR | 05119002800 | 51.30 | 46.70 | 28.80 | 18.00 | 2.64 |
| 20 | Webster | LA | 22119031700 | 52.70 | 54.30 | 25.80 | 0.00 | 2.51 |
| 21 | Navajo | AZ | 04017940014 | 56.40 | 66.70 | 22.40 | 1.83 | 2.17 |
| 22 | Mobile | AL | 01097001501 | 44.90 | 47.50 | 21.60 | 9.23 | 3.15 |
| 23 | Calhoun | AL | 01015000500 | 50.60 | 45.00 | 29.00 | 4.29 | 2.64 |
| 24 | Harrison | MS | 28047001800 | 48.30 | 44.40 | 27.00 | 4.27 | 2.73 |
| 25 | Lauderdale | MS | 28075010700 | 49.60 | 44.60 | 28.20 | 8.51 | 2.59 |

**Table S2.** Twenty-five highest-ranked deployment-priority census tracts. Tracts are ranked by the deployment-priority score, calculated as standardized burden minus standardized DDI. Other definitions are as in Table S1.

| Rank | County | State | Tract FIPS | DDI composite | INFA | SE | Cardiologists per 100,000 | Burden z score |
| --- | --- | --- | --- | --- | --- | --- | --- | --- |
| 1 | Marion | IN | 18097357602 | 15.00 | 7.80 | 13.60 | 9.19 | 1.45 |
| 2 | Chatham | GA | 13051003302 | 16.00 | 19.50 | 7.80 | 8.89 | 1.46 |
| 3 | Henrico | VA | 51087201405 | 17.30 | 13.20 | 12.90 | 9.86 | 1.49 |
| 4 | Mobile | AL | 01097005100 | 13.50 | 11.90 | 9.60 | 9.23 | 1.14 |
| 5 | Milwaukee | WI | 55079004000 | 17.10 | 20.40 | 8.40 | 8.51 | 1.45 |
| 6 | Marion | FL | 12083001206 | 18.40 | 16.80 | 11.90 | 6.10 | 1.53 |
| 7 | Miami-Dade | FL | 12086014100 | 17.70 | 8.80 | 15.80 | 10.98 | 1.42 |
| 8 | Harris | TX | 48201331603 | 18.50 | 12.80 | 14.30 | 7.36 | 1.48 |
| 9 | Clayton | GA | 13063040536 | 11.60 | 9.10 | 9.30 | 5.03 | 0.87 |
| 10 | Tunica | MS | 28143950102 | 18.30 | 19.00 | 10.50 | 0.00 | 1.45 |
| 11 | Wayne | MS | 28153950301 | 14.80 | 9.60 | 12.40 | 0.00 | 1.11 |
| 12 | Marion | IN | 18097381207 | 11.80 | 9.80 | 9.10 | 9.19 | 0.84 |
| 13 | Marion | IN | 18097340600 | 17.90 | 15.00 | 12.40 | 9.19 | 1.38 |
| 14 | Harris | TX | 48201453702 | 15.10 | 13.00 | 10.60 | 7.36 | 1.08 |
| 15 | St. Louis | MO | 29189210501 | 18.00 | 18.30 | 10.60 | 14.99 | 1.32 |
| 16 | Prince George's | MD | 24033803001 | 14.90 | 14.50 | 9.60 | 3.59 | 1.02 |
| 17 | Marion | IN | 18097310306 | 16.30 | 18.40 | 8.70 | 9.19 | 1.14 |
| 18 | Cabarrus | NC | 37025040705 | 7.50 | 8.20 | 5.50 | 2.08 | 0.35 |
| 19 | Lee | AL | 01081041702 | 17.00 | 10.90 | 13.90 | 4.91 | 1.19 |
| 20 | Harris | TX | 48201550305 | 8.20 | 9.40 | 5.60 | 7.36 | 0.41 |
| 21 | Honolulu | HI | 15003009803 | 16.40 | 13.40 | 11.80 | 4.55 | 1.14 |
| 22 | Blount | TN | 47009010100 | 16.10 | 11.20 | 12.70 | 2.83 | 1.10 |
| 23 | Hernando | FL | 12053041602 | 18.70 | 13.50 | 14.10 | 4.23 | 1.33 |
| 24 | Broward | FL | 12011060302 | 16.60 | 18.50 | 9.00 | 9.12 | 1.15 |
| 25 | Mobile | AL | 01097006903 | 15.40 | 10.60 | 12.40 | 9.23 | 1.01 |

**Table S3.** Cardiometabolic burden of the 19 Delaware County, Pennsylvania tracts that appeared in the primary top-25 deployment list, under the primary single-measure composite versus a full-measure composite rebuilt from the last release in which Pennsylvania reported all 10 measures and crosswalked onto current tracts. Sixteen of the 19 tracts remained above the national-average burden.

| Rank | Tract FIPS | Sleep-only burden (z) | Full-measure burden (z) | Above-average burden |
| --- | --- | --- | --- | --- |
| 1 | 42045404900 | 3.40 | 1.70 | Yes |
| 2 | 42045406401 | 2.91 | 0.71 | Yes |
| 3 | 42045402700 | 3.22 | 0.13 | Yes |
| 4 | 42045402600 | 3.04 | 0.63 | Yes |
| 5 | 42045404800 | 2.94 | 1.15 | Yes |
| 6 | 42045402100 | 2.68 | 0.52 | Yes |
| 7 | 42045400301 | 2.87 | 0.39 | Yes |
| 8 | 42045401700 | 2.67 | 0.24 | Yes |
| 9 | 42045400402 | 2.72 | 0.65 | Yes |
| 10 | 42045404600 | 2.52 | 0.39 | Yes |
| 11 | 42045403101 | 2.46 | 0.11 | Yes |
| 12 | 42045401800 | 1.85 | -0.04 | No |
| 13 | 42045401402 | 1.72 | -0.08 | No |
| 14 | 42045403401 | 2.00 | 0.17 | Yes |
| 15 | 42045404400 | 2.11 | 0.40 | Yes |
| 16 | 42045406600 | 2.08 | 0.66 | Yes |
| 17 | 42045400500 | 2.08 | 0.01 | Yes |
| 18 | 42045401900 | 1.58 | 0.10 | Yes |
| 19 | 42045401401 | 1.15 | -0.54 | No |

**Table S4.** Top 25 investment tracts with readiness measured by the raw components of the Divide Index, 6 American Community Survey indicators plus FCC terrestrial broadband availability, combined as an equal-weight z score mean. The final column indicates whether each tract also appears in the primary top 25 (Table S1).

| Rank | County | State | Tract FIPS | Components index (z) | DDI composite | Burden z score | In primary top 25 |
| --- | --- | --- | --- | --- | --- | --- | --- |
| 1 | Socorro | NM | 35053940000 | 7.86 | 70.00 | 2.06 | Yes |
| 2 | Orange | FL | 12095010500 | 4.17 | 78.80 | 2.79 | Yes |
| 3 | Bernalillo | NM | 35001940602 | 5.05 | 46.60 | 1.28 | No |
| 4 | Orleans | LA | 22071000620 | 3.47 | 56.80 | 2.69 | Yes |
| 5 | Navajo | AZ | 04017942400 | 3.97 | 62.60 | 1.90 | Yes |
| 6 | Lake | IN | 18089011700 | 3.26 | 70.80 | 2.68 | Yes |
| 7 | Cook | IL | 17031461000 | 3.40 | 48.70 | 2.46 | No |
| 8 | Mahoning | OH | 39099802100 | 2.81 | 37.10 | 2.89 | No |
| 9 | Navajo | AZ | 04017940008 | 3.58 | 50.50 | 1.95 | No |
| 10 | Harris | TX | 48201331400 | 2.25 | 47.20 | 3.41 | Yes |
| 11 | Sandoval | NM | 35043940900 | 3.60 | 60.90 | 1.81 | Yes |
| 12 | Lauderdale | MS | 28075010700 | 2.84 | 49.60 | 2.59 | Yes |
| 13 | Lake | IN | 18089011900 | 2.35 | 46.80 | 3.06 | Yes |
| 14 | Navajo | AZ | 04017940010 | 3.26 | 52.00 | 2.00 | No |
| 15 | Westchester | NY | 36119984000 | 2.39 | 40.90 | 2.91 | No |
| 16 | Navajo | AZ | 04017940014 | 3.04 | 56.40 | 2.17 | Yes |
| 17 | Dougherty | GA | 13095011400 | 2.85 | 44.70 | 2.35 | No |
| 18 | Lowndes | AL | 01085781100 | 2.34 | 36.60 | 2.87 | No |
| 19 | Orleans | LA | 22071001751 | 2.61 | 56.60 | 2.53 | Yes |
| 20 | Lauderdale | MS | 28075000401 | 2.94 | 38.00 | 2.14 | No |
| 21 | Greene | AL | 01063060200 | 3.11 | 45.80 | 1.93 | No |
| 22 | Ouachita | LA | 22073010700 | 2.03 | 47.80 | 3.15 | Yes |
| 23 | Greene | AL | 01063060000 | 2.02 | 48.90 | 3.12 | Yes |
| 24 | Norfolk City | VA | 51710004200 | 2.10 | 40.30 | 3.02 | No |
| 25 | San Juan | NM | 35045942900 | 3.55 | 55.20 | 1.35 | No |

**Table S5.** Top 25 deployment tracts with readiness measured by the raw components of the Divide Index. Columns follow Table S4, with overlap against the primary top 25 marked in the final column (Table S2).

| Rank | County | State | Tract FIPS | Components index (z) | DDI composite | Burden z score | In primary top 25 |
| --- | --- | --- | --- | --- | --- | --- | --- |
| 1 | Ouachita | LA | 22073010103 | -0.36 | 20.60 | 1.79 | No |
| 2 | Oklahoma | OK | 40109109800 | -0.11 | 23.40 | 1.96 | No |
| 3 | Milwaukee | WI | 55079004200 | -0.10 | 27.70 | 1.86 | No |
| 4 | Suffolk City | VA | 51800065100 | -0.26 | 24.40 | 1.48 | No |
| 5 | Marion | FL | 12083001206 | -0.16 | 18.40 | 1.53 | Yes |
| 6 | Hamilton | OH | 39061021602 | -0.72 | 15.50 | 0.87 | No |
| 7 | Milwaukee | WI | 55079004000 | -0.21 | 17.10 | 1.45 | Yes |
| 8 | Pike | MS | 28113950103 | -0.10 | 31.80 | 1.56 | No |
| 9 | St. Tammany | LA | 22103041106 | -0.29 | 21.00 | 1.31 | No |
| 10 | Wood | WV | 54107000801 | -0.15 | 19.60 | 1.41 | No |
| 11 | Fort Bend | TX | 48157670300 | -0.19 | 20.20 | 1.30 | No |
| 12 | Orange | FL | 12095012304 | -0.49 | 23.20 | 0.92 | No |
| 13 | Harris | TX | 48201453702 | -0.35 | 15.10 | 1.08 | Yes |
| 14 | Jefferson | TX | 48245010200 | -0.36 | 16.60 | 1.06 | No |
| 15 | Broward | FL | 12011060302 | -0.28 | 16.60 | 1.15 | Yes |
| 16 | Clayton | GA | 13063040624 | -0.38 | 16.90 | 1.03 | No |
| 17 | Prince George's | MD | 24033803001 | -0.37 | 14.90 | 1.02 | Yes |
| 18 | Logan | OK | 40083600300 | -0.38 | 16.80 | 1.00 | No |
| 19 | Polk | FL | 12105014128 | -0.50 | 13.90 | 0.85 | No |
| 20 | Clayton | GA | 13063040615 | -0.59 | 15.70 | 0.74 | No |
| 21 | Hampton City | VA | 51650012000 | -0.18 | 18.20 | 1.21 | No |
| 22 | Vermilion | IL | 17183000200 | -0.18 | 22.30 | 1.21 | No |
| 23 | Essex | NJ | 34013004100 | -0.57 | 20.50 | 0.75 | No |
| 24 | Lafourche | LA | 22057021903 | -0.20 | 20.30 | 1.16 | No |
| 25 | Kankakee | IL | 17091011600 | -0.13 | 24.30 | 1.23 | No |

**Table S6.** Top 25 investment tracts with readiness measured by the infrastructure/adoption (INFA) subscore alone, anchored at its national median of 17.5. Tracts also present in the primary top 25 are flagged in the final column (Table S1).

| Rank | County | State | Tract FIPS | INFA score | DDI composite | Burden z score | In primary top 25 |
| --- | --- | --- | --- | --- | --- | --- | --- |
| 1 | Apache | AZ | 04001942600 | 100.00 | 67.40 | 1.25 | Yes |
| 2 | Socorro | NM | 35053940000 | 81.70 | 70.00 | 2.06 | Yes |
| 3 | Navajo | AZ | 04017942400 | 83.30 | 62.60 | 1.90 | Yes |
| 4 | Orange | FL | 12095010500 | 71.20 | 78.80 | 2.79 | Yes |
| 5 | Lake | IN | 18089011700 | 69.20 | 70.80 | 2.68 | Yes |
| 6 | Apache | AZ | 04001944301 | 83.30 | 58.20 | 1.23 | No |
| 7 | Sandoval | NM | 35043940900 | 73.90 | 60.90 | 1.81 | Yes |
| 8 | Orleans | LA | 22071000620 | 63.00 | 56.80 | 2.69 | Yes |
| 9 | Apache | AZ | 04001944302 | 76.10 | 54.40 | 1.41 | No |
| 10 | Navajo | AZ | 04017940014 | 66.70 | 56.40 | 2.17 | Yes |
| 11 | Shelby | TN | 47157005800 | 55.30 | 51.90 | 3.06 | Yes |
| 12 | Shelby | TN | 47157005700 | 61.20 | 43.80 | 2.54 | No |
| 13 | Shelby | TN | 47157005500 | 59.50 | 54.20 | 2.63 | Yes |
| 14 | Shelby | TN | 47157005900 | 52.00 | 53.40 | 3.24 | Yes |
| 15 | Ouachita | LA | 22073010700 | 52.70 | 47.80 | 3.15 | Yes |
| 16 | Apache | AZ | 04001945002 | 72.40 | 51.30 | 1.31 | No |
| 17 | San Juan | NM | 35045942900 | 71.80 | 55.20 | 1.35 | No |
| 18 | Lake | IN | 18089011900 | 51.80 | 46.80 | 3.06 | Yes |
| 19 | Webster | LA | 22119031900 | 50.40 | 47.70 | 3.17 | Yes |
| 20 | Shelby | TN | 47157000600 | 58.70 | 57.90 | 2.45 | Yes |
| 21 | Apache | AZ | 04001944202 | 76.30 | 52.30 | 0.90 | No |
| 22 | Greene | AL | 01063060000 | 50.30 | 48.90 | 3.12 | Yes |
| 23 | Monroe | AL | 01099075800 | 65.40 | 45.80 | 1.80 | No |
| 24 | San Juan | NM | 35045943100 | 70.30 | 53.10 | 1.34 | No |
| 25 | Laurens | GA | 13175950900 | 56.70 | 44.20 | 2.51 | No |

**Table S7.** Top 25 deployment tracts with readiness measured by the INFA subscore alone. Columns follow Table S6, and the final column records overlap with the primary top 25 (Table S2).

| Rank | County | State | Tract FIPS | INFA score | DDI composite | Burden z score | In primary top 25 |
| --- | --- | --- | --- | --- | --- | --- | --- |
| 1 | Westchester | NY | 36119984000 | 12.70 | 40.90 | 2.91 | No |
| 2 | Cook | IL | 17031670700 | 12.50 | 23.50 | 2.21 | No |
| 3 | Harris | TX | 48201231500 | 11.30 | 19.00 | 1.90 | No |
| 4 | Marion | IN | 18097357602 | 7.80 | 15.00 | 1.45 | Yes |
| 5 | Harris | TX | 48201232000 | 9.40 | 20.50 | 1.50 | No |
| 6 | Miami-Dade | FL | 12086014100 | 8.80 | 17.70 | 1.42 | Yes |
| 7 | Hamilton | OH | 39061007700 | 15.60 | 22.70 | 1.97 | No |
| 8 | Montgomery | OH | 39113004200 | 10.40 | 20.00 | 1.47 | No |
| 9 | Harris | TX | 48201231900 | 13.70 | 24.50 | 1.74 | No |
| 10 | Shelby | TN | 47157010002 | 14.10 | 22.50 | 1.76 | No |
| 11 | Hudson | NJ | 34017006900 | 8.10 | 24.40 | 1.23 | No |
| 12 | Fulton | GA | 13121004800 | 14.40 | 25.10 | 1.73 | No |
| 13 | Orleans | LA | 22071000903 | 16.90 | 28.90 | 1.89 | No |
| 14 | Ouachita | LA | 22073010103 | 15.80 | 20.60 | 1.79 | No |
| 15 | Harris | TX | 48201231400 | 11.70 | 21.70 | 1.39 | No |
| 16 | Brevard | FL | 12009069906 | 13.60 | 24.30 | 1.56 | No |
| 17 | Harris | TX | 48201331603 | 12.80 | 18.50 | 1.48 | Yes |
| 18 | Harris | TX | 48201231100 | 9.60 | 21.30 | 1.19 | No |
| 19 | Henrico | VA | 51087201405 | 13.20 | 17.30 | 1.49 | Yes |
| 20 | Harrison | MS | 28047001900 | 13.50 | 28.20 | 1.51 | No |
| 21 | Harris | TX | 48201231600 | 13.20 | 28.10 | 1.46 | No |
| 22 | Harris | TX | 48201453902 | 10.40 | 18.50 | 1.20 | No |
| 23 | Wood | WV | 54107000702 | 15.50 | 19.60 | 1.63 | No |
| 24 | Wayne | MS | 28153950301 | 9.60 | 14.80 | 1.11 | Yes |
| 25 | St. Louis | MO | 29189212102 | 17.40 | 24.00 | 1.77 | No |

**Table S8.** Top 25 investment tracts with readiness measured by the socioeconomic (SE) subscore alone, anchored at its national median of 11.5. The final column identifies tracts shared with the primary top 25 (Table S1).

| Rank | County | State | Tract FIPS | SE score | DDI composite | Burden z score | In primary top 25 |
| --- | --- | --- | --- | --- | --- | --- | --- |
| 1 | Orange | FL | 12095010500 | 43.10 | 78.80 | 2.79 | Yes |
| 2 | Jackson | TN | 47087960300 | 53.60 | 62.60 | 0.74 | No |
| 3 | Westchester | NY | 36119984000 | 37.80 | 40.90 | 2.91 | No |
| 4 | Lake | IN | 18089011700 | 36.00 | 70.80 | 2.68 | Yes |
| 5 | Harris | TX | 48201331400 | 30.80 | 47.20 | 3.41 | Yes |
| 6 | Cook | IL | 17031835600 | 35.80 | 50.40 | 2.48 | No |
| 7 | Harris | TX | 48201422405 | 35.30 | 49.70 | 2.32 | No |
| 8 | Westchester | NY | 36119000502 | 35.20 | 52.00 | 2.29 | No |
| 9 | Mercer | NJ | 34021000100 | 46.80 | 63.40 | 0.44 | No |
| 10 | Shelby | TN | 47157005900 | 27.90 | 53.40 | 3.24 | Yes |
| 11 | Morgan | MO | 29141470502 | 31.40 | 40.90 | 2.63 | No |
| 12 | McDowell | WV | 54047953900 | 33.00 | 45.70 | 2.31 | No |
| 13 | Orleans | LA | 22071001751 | 31.40 | 56.60 | 2.53 | Yes |
| 14 | Montgomery | AL | 01101002400 | 29.50 | 53.70 | 2.77 | Yes |
| 15 | Lafayette | LA | 22055000900 | 25.50 | 39.70 | 3.29 | No |
| 16 | Hamilton | OH | 39061026300 | 29.00 | 44.20 | 2.76 | No |
| 17 | Cook | IL | 17031260400 | 33.60 | 44.90 | 1.96 | No |
| 18 | Calhoun | AL | 01015000500 | 29.00 | 50.60 | 2.64 | Yes |
| 19 | Iberia | LA | 22045030202 | 41.80 | 50.10 | 0.65 | No |
| 20 | Montgomery | AL | 01101001000 | 31.70 | 47.20 | 2.20 | No |
| 21 | Norfolk City | VA | 51710004200 | 26.40 | 40.30 | 3.02 | No |
| 22 | Montgomery | AL | 01101001200 | 29.70 | 45.50 | 2.50 | No |
| 23 | Pulaski | AR | 05119002800 | 28.80 | 51.30 | 2.64 | Yes |
| 24 | Acadia | LA | 22001961000 | 32.60 | 54.30 | 1.97 | No |
| 25 | Mahoning | OH | 39099800501 | 27.40 | 46.00 | 2.72 | No |

**Table S9.** Top 25 deployment tracts with readiness measured by the SE subscore alone. Columns follow Table S8; overlap with the primary top 25 is given in the final column (Table S2).

| Rank | County | State | Tract FIPS | SE score | DDI composite | Burden z score | In primary top 25 |
| --- | --- | --- | --- | --- | --- | --- | --- |
| 1 | Lake | IN | 18089011600 | 11.20 | 19.70 | 2.30 | No |
| 2 | Bullock | AL | 01011952100 | 11.30 | 30.40 | 2.18 | No |
| 3 | Clark | OH | 39023000902 | 11.50 | 25.60 | 2.06 | No |
| 4 | Chatham | GA | 13051003302 | 7.80 | 16.00 | 1.46 | Yes |
| 5 | Milwaukee | WI | 55079004000 | 8.40 | 17.10 | 1.45 | Yes |
| 6 | Menominee | WI | 55078940101 | 9.30 | 21.50 | 1.49 | No |
| 7 | Milwaukee | WI | 55079002500 | 10.80 | 21.50 | 1.60 | No |
| 8 | Tunica | MS | 28143950102 | 10.50 | 18.30 | 1.45 | Yes |
| 9 | Marion | IN | 18097310306 | 8.70 | 16.30 | 1.14 | Yes |
| 10 | Shelby | TN | 47157022321 | 11.30 | 30.90 | 1.51 | No |
| 11 | Broward | FL | 12011060302 | 9.00 | 16.60 | 1.15 | Yes |
| 12 | Muskegon | MI | 26121001200 | 11.10 | 21.80 | 1.47 | No |
| 13 | Laurens | GA | 13175951002 | 9.90 | 23.70 | 1.28 | No |
| 14 | Fulton | GA | 13121010604 | 11.00 | 22.10 | 1.45 | No |
| 15 | Hamilton | OH | 39061021602 | 7.40 | 15.50 | 0.87 | No |
| 16 | St. Landry | LA | 22097960200 | 10.60 | 27.70 | 1.37 | No |
| 17 | Mobile | AL | 01097002400 | 10.50 | 21.30 | 1.35 | No |
| 18 | St. James | LA | 22093040400 | 7.00 | 17.80 | 0.78 | No |
| 19 | Kankakee | IL | 17091011600 | 9.90 | 24.30 | 1.23 | No |
| 20 | Gaston | NC | 37071031800 | 11.40 | 25.60 | 1.45 | No |
| 21 | St. Louis | MO | 29189210501 | 10.60 | 18.00 | 1.32 | Yes |
| 22 | Suffolk City | VA | 51800065402 | 9.70 | 29.90 | 1.16 | No |
| 23 | Charleston | SC | 45019002402 | 9.90 | 24.40 | 1.19 | No |
| 24 | Mobile | AL | 01097005100 | 9.60 | 13.50 | 1.14 | Yes |
| 25 | Hardee | FL | 12049970202 | 8.00 | 17.50 | 0.88 | No |

**Table S10.** Top 25 investment tracts when the matrix covers all US tracts without the workforce screen. The final column flags whether the tract lies in a workforce-constrained county, one with no cardiologists or a declining per-capita cardiology workforce. Tracts in the 9800 series, which the Census Bureau reserves for special land use including institutional and port areas, are retained; the highest-ranked tract, 06037980014, comprises the Terminal Island federal correctional and port complex in Los Angeles County.

| Rank | County | State | Tract FIPS | DDI composite | Burden z score | In workforce-constrained pool |
| --- | --- | --- | --- | --- | --- | --- |
| 1 | Los Angeles | CA | 06037980014 | 78.70 | 3.83 | No |
| 2 | Orange | FL | 12095010500 | 78.80 | 2.79 | Yes |
| 3 | Bibb | GA | 13021011500 | 70.80 | 3.40 | No |
| 4 | Kings | NY | 36047035200 | 77.40 | 2.17 | No |
| 5 | Lake | IN | 18089011700 | 70.80 | 2.68 | Yes |
| 6 | Los Angeles | CA | 06037206301 | 64.10 | 3.12 | No |
| 7 | Wayne | MI | 26163559800 | 64.30 | 2.88 | No |
| 8 | Washington | MS | 28151000400 | 61.20 | 2.93 | No |
| 9 | Cuyahoga | OH | 39035152701 | 57.70 | 3.19 | No |
| 10 | Socorro | NM | 35053940000 | 70.00 | 2.06 | Yes |
| 11 | Cuyahoga | OH | 39035109301 | 57.30 | 3.20 | No |
| 12 | El Paso | TX | 48141001800 | 57.40 | 3.14 | No |
| 13 | Cuyahoga | OH | 39035112100 | 49.20 | 3.80 | No |
| 14 | New York | NY | 36061011900 | 69.30 | 1.96 | No |
| 15 | Shelby | TN | 47157005900 | 53.40 | 3.24 | Yes |
| 16 | McKinley | NM | 35031945702 | 62.80 | 2.35 | No |
| 17 | Caddo | LA | 22017025200 | 62.50 | 2.36 | No |
| 18 | Cuyahoga | OH | 39035117201 | 52.90 | 3.21 | No |
| 19 | Dallas | AL | 01047957100 | 65.00 | 2.05 | No |
| 20 | Orleans | LA | 22071000620 | 56.80 | 2.69 | Yes |
| 21 | Shelby | TN | 47157005800 | 51.90 | 3.06 | Yes |
| 22 | Shelby | TN | 47157000600 | 57.90 | 2.45 | Yes |
| 23 | Harris | TX | 48201331400 | 47.20 | 3.41 | Yes |
| 24 | Orleans | LA | 22071001751 | 56.60 | 2.53 | Yes |
| 25 | Caddo | LA | 22017020600 | 51.80 | 2.96 | No |

**Table S11.** Top 25 deployment tracts when the matrix covers all US tracts without the workforce screen. Columns follow Table S10.

| Rank | County | State | Tract FIPS | DDI composite | Burden z score | In workforce-constrained pool |
| --- | --- | --- | --- | --- | --- | --- |
| 1 | Queens | NY | 36081053902 | 2.60 | 0.53 | No |
| 2 | South Central CT PR | CT | 09170142605 | 14.20 | 1.55 | No |
| 3 | Hinds | MS | 28049011002 | 9.80 | 1.14 | No |
| 4 | Naugatuck Valley PR | CT | 09140350200 | 14.80 | 1.46 | No |
| 5 | Horry | SC | 45051050902 | 14.50 | 1.44 | No |
| 6 | Marion | IN | 18097357602 | 15.00 | 1.45 | Yes |
| 7 | Jefferson | AL | 01073011806 | 16.10 | 1.53 | No |
| 8 | Chatham | GA | 13051003302 | 16.00 | 1.46 | Yes |
| 9 | Dallas | TX | 48113016526 | 10.80 | 0.92 | No |
| 10 | Henrico | VA | 51087201405 | 17.30 | 1.49 | Yes |
| 11 | Mobile | AL | 01097005100 | 13.50 | 1.14 | Yes |
| 12 | South Central CT PR | CT | 09170140600 | 13.00 | 1.08 | No |
| 13 | Milwaukee | WI | 55079004000 | 17.10 | 1.45 | Yes |
| 14 | Marion | FL | 12083001206 | 18.40 | 1.53 | Yes |
| 15 | Wayne | MI | 26163538800 | 13.10 | 1.05 | No |
| 16 | DeKalb | GA | 13089023331 | 13.30 | 1.03 | No |
| 17 | Clayton | GA | 13063040536 | 11.60 | 0.87 | Yes |
| 18 | Miami-Dade | FL | 12086014100 | 17.70 | 1.42 | Yes |
| 19 | Harris | TX | 48201331603 | 18.50 | 1.48 | Yes |
| 20 | Tunica | MS | 28143950102 | 18.30 | 1.45 | Yes |
| 21 | Greater Bridgeport PR | CT | 09120257200 | 7.00 | 0.41 | No |
| 22 | Marion | IN | 18097381207 | 11.80 | 0.84 | Yes |
| 23 | Wayne | MS | 28153950301 | 14.80 | 1.11 | Yes |
| 24 | Marion | IN | 18097340600 | 17.90 | 1.38 | Yes |
| 25 | Wayne | GA | 13305970201 | 16.10 | 1.22 | No |

**Table S12.** Box sizes and survival of the primary top-10 lists under stricter national classification thresholds. Each row applies one combination of burden and readiness cutoffs. Every stricter box is a subset of the primary box, so surviving tracts keep their primary rank.

| Burden cutoff | Readiness cutoff | Investment tracts | Deployment tracts | Primary top-10 investment retained | Primary top-10 deployment retained |
| --- | --- | --- | --- | --- | --- |
| Above mean ($z > 0$) | Median split (18.8) | 21,752 | 4,765 | 10/10 | 10/10 |
| Above mean ($z > 0$) | Quartile split (14.2 / 24.2) | 13,095 | 855 | 10/10 | 0/10 |
| Above mean ($z > 0$) | Decile split (10.9 / 29.8) | 5,484 | 114 | 10/10 | 0/10 |
| Top 25% ($z \geq 0.43$) | Median split (18.8) | 13,261 | 1,000 | 10/10 | 10/10 |
| Top 25% ($z \geq 0.43$) | Quartile split (14.2 / 24.2) | 9,879 | 133 | 10/10 | 0/10 |
| Top 25% ($z \geq 0.43$) | Decile split (10.9 / 29.8) | 4,818 | 15 | 10/10 | 0/10 |
| Top 10% ($z \geq 0.91$) | Median split (18.8) | 5,334 | 174 | 10/10 | 10/10 |
| Top 10% ($z \geq 0.91$) | Quartile split (14.2 / 24.2) | 4,674 | 29 | 10/10 | 0/10 |
| Top 10% ($z \geq 0.91$) | Decile split (10.9 / 29.8) | 2,971 | 8 | 10/10 | 0/10 |

**Table S13.** Top 25 investment tracts with the workforce-constrained pool redefined by 25-mile cross-county access, counties with zero cardiologists within 25 miles of the county centroid or a declining per-capita rate of the aggregated workforce. Tracts common to the primary top 25 are indicated in the final column (Table S1).

| Rank | County | State | Tract FIPS | DDI composite | Burden z score | In primary top 25 |
| --- | --- | --- | --- | --- | --- | --- |
| 1 | Orange | FL | 12095010500 | 78.80 | 2.79 | Yes |
| 2 | Bibb | GA | 13021011500 | 70.80 | 3.40 | No |
| 3 | Lake | IN | 18089011700 | 70.80 | 2.68 | Yes |
| 4 | Socorro | NM | 35053940000 | 70.00 | 2.06 | Yes |
| 5 | Shelby | TN | 47157005900 | 53.40 | 3.24 | Yes |
| 6 | Orleans | LA | 22071000620 | 56.80 | 2.69 | Yes |
| 7 | Shelby | TN | 47157005800 | 51.90 | 3.06 | Yes |
| 8 | Shelby | TN | 47157000600 | 57.90 | 2.45 | Yes |
| 9 | Harris | TX | 48201331400 | 47.20 | 3.41 | Yes |
| 10 | Orleans | LA | 22071001751 | 56.60 | 2.53 | Yes |
| 11 | Montgomery | AL | 01101002400 | 53.70 | 2.77 | Yes |
| 12 | Navajo | AZ | 04017942400 | 62.60 | 1.90 | Yes |
| 13 | Duval | FL | 12031001000 | 55.40 | 2.55 | No |
| 14 | Greene | AL | 01063060000 | 48.90 | 3.12 | Yes |
| 15 | Shelby | TN | 47157005500 | 54.20 | 2.63 | Yes |
| 16 | Ouachita | LA | 22073010700 | 47.80 | 3.15 | Yes |
| 17 | Apache | AZ | 04001942600 | 67.40 | 1.25 | Yes |
| 18 | Sandoval | NM | 35043940900 | 60.90 | 1.81 | Yes |
| 19 | Lake | IN | 18089011900 | 46.80 | 3.06 | Yes |
| 20 | Pulaski | AR | 05119002800 | 51.30 | 2.64 | Yes |
| 21 | Navajo | AZ | 04017940014 | 56.40 | 2.17 | Yes |
| 22 | Calhoun | AL | 01015000500 | 50.60 | 2.64 | Yes |
| 23 | Mobile | AL | 01097001501 | 44.90 | 3.15 | Yes |
| 24 | Harrison | MS | 28047001800 | 48.30 | 2.73 | Yes |
| 25 | Lauderdale | MS | 28075010700 | 49.60 | 2.59 | Yes |

**Table S14.** Top 25 deployment tracts with the workforce-constrained pool redefined by 25-mile cross-county access. Columns follow Table S13, and the final column records overlap with the primary top 25 (Table S2).

| Rank | County | State | Tract FIPS | DDI composite | Burden z score | In primary top 25 |
| --- | --- | --- | --- | --- | --- | --- |
| 1 | Marion | IN | 18097357602 | 15.00 | 1.45 | Yes |
| 2 | Jefferson | AL | 01073011806 | 16.10 | 1.53 | No |
| 3 | Chatham | GA | 13051003302 | 16.00 | 1.46 | Yes |
| 4 | Henrico | VA | 51087201405 | 17.30 | 1.49 | Yes |
| 5 | Mobile | AL | 01097005100 | 13.50 | 1.14 | Yes |
| 6 | Milwaukee | WI | 55079004000 | 17.10 | 1.45 | Yes |
| 7 | Marion | FL | 12083001206 | 18.40 | 1.53 | Yes |
| 8 | Miami-Dade | FL | 12086014100 | 17.70 | 1.42 | Yes |
| 9 | Harris | TX | 48201331603 | 18.50 | 1.48 | Yes |
| 10 | Tunica | MS | 28143950102 | 18.30 | 1.45 | Yes |
| 11 | Marion | IN | 18097381207 | 11.80 | 0.84 | Yes |
| 12 | Wayne | MS | 28153950301 | 14.80 | 1.11 | Yes |
| 13 | Marion | IN | 18097340600 | 17.90 | 1.38 | Yes |
| 14 | Harris | TX | 48201453702 | 15.10 | 1.08 | Yes |
| 15 | Livingston | LA | 22063040503 | 12.40 | 0.81 | No |
| 16 | St. Louis | MO | 29189210501 | 18.00 | 1.32 | Yes |
| 17 | Cabarrus | NC | 37025040705 | 7.50 | 0.35 | Yes |
| 18 | Harris | TX | 48201550305 | 8.20 | 0.41 | Yes |
| 19 | Marion | IN | 18097310306 | 16.30 | 1.14 | Yes |
| 20 | Lee | AL | 01081041702 | 17.00 | 1.19 | Yes |
| 21 | Honolulu | HI | 15003009803 | 16.40 | 1.14 | Yes |
| 22 | Blount | TN | 47009010100 | 16.10 | 1.10 | Yes |
| 23 | Broward | FL | 12011060302 | 16.60 | 1.15 | Yes |
| 24 | Hernando | FL | 12053041602 | 18.70 | 1.33 | Yes |
| 25 | Comal | TX | 48091310612 | 17.50 | 1.22 | No |

**Table S15.**
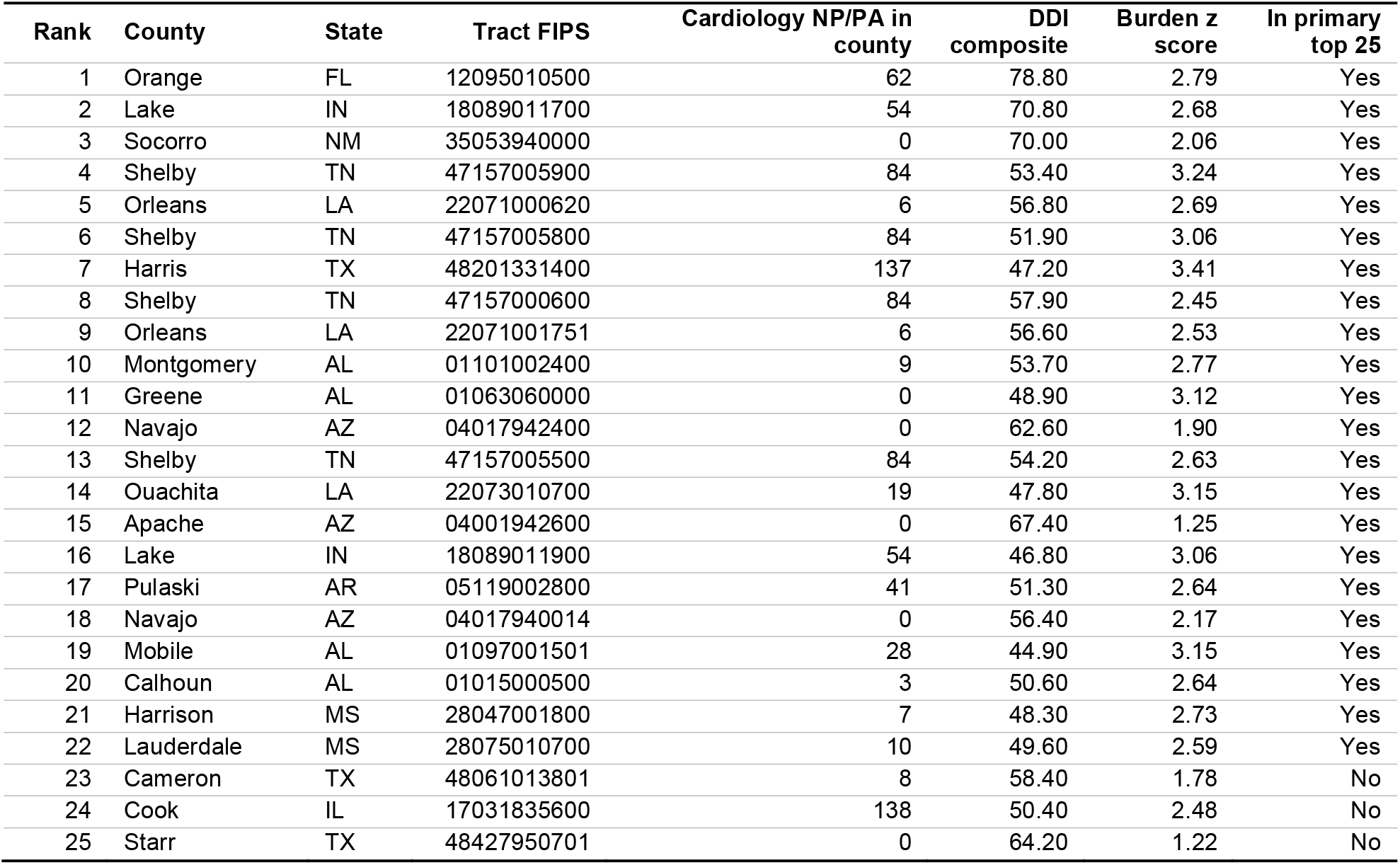
Top 25 investment tracts with the workforce-constrained pool redefined by the generalized cardiology workforce, cardiologists plus cardiology nurse practitioners and physician assistants attributed through cardiology-majority practice sites in the CMS Doctors and Clinicians file. The NP/PA column counts site-strict cardiology NPs and PAs in the tract’s parent county. The final column shows which tracts also appear in the primary top 25 (Table S1).

| Rank | County | State | Tract FIPS | Cardiology NP/PA in county | DDI composite | Burden z score | In primary top 25 |
| --- | --- | --- | --- | --- | --- | --- | --- |
| 1 | Orange | FL | 12095010500 | 62 | 78.80 | 2.79 | Yes |
| 2 | Lake | IN | 18089011700 | 54 | 70.80 | 2.68 | Yes |
| 3 | Socorro | NM | 35053940000 | 0 | 70.00 | 2.06 | Yes |
| 4 | Shelby | TN | 47157005900 | 84 | 53.40 | 3.24 | Yes |
| 5 | Orleans | LA | 22071000620 | 6 | 56.80 | 2.69 | Yes |
| 6 | Shelby | TN | 47157005800 | 84 | 51.90 | 3.06 | Yes |
| 7 | Harris | TX | 48201331400 | 137 | 47.20 | 3.41 | Yes |
| 8 | Shelby | TN | 47157000600 | 84 | 57.90 | 2.45 | Yes |
| 9 | Orleans | LA | 22071001751 | 6 | 56.60 | 2.53 | Yes |
| 10 | Montgomery | AL | 01101002400 | 9 | 53.70 | 2.77 | Yes |
| 11 | Greene | AL | 01063060000 | 0 | 48.90 | 3.12 | Yes |
| 12 | Navajo | AZ | 04017942400 | 0 | 62.60 | 1.90 | Yes |
| 13 | Shelby | TN | 47157005500 | 84 | 54.20 | 2.63 | Yes |
| 14 | Ouachita | LA | 22073010700 | 19 | 47.80 | 3.15 | Yes |
| 15 | Apache | AZ | 04001942600 | 0 | 67.40 | 1.25 | Yes |
| 16 | Lake | IN | 18089011900 | 54 | 46.80 | 3.06 | Yes |
| 17 | Pulaski | AR | 05119002800 | 41 | 51.30 | 2.64 | Yes |
| 18 | Navajo | AZ | 04017940014 | 0 | 56.40 | 2.17 | Yes |
| 19 | Mobile | AL | 01097001501 | 28 | 44.90 | 3.15 | Yes |
| 20 | Calhoun | AL | 01015000500 | 3 | 50.60 | 2.64 | Yes |
| 21 | Harrison | MS | 28047001800 | 7 | 48.30 | 2.73 | Yes |
| 22 | Lauderdale | MS | 28075010700 | 10 | 49.60 | 2.59 | Yes |
| 23 | Cameron | TX | 48061013801 | 8 | 58.40 | 1.78 | No |
| 24 | Cook | IL | 17031835600 | 138 | 50.40 | 2.48 | No |
| 25 | Starr | TX | 48427950701 | 0 | 64.20 | 1.22 | No |

**Table S16.** Top 25 deployment tracts with the workforce-constrained pool redefined by the generalized cardiology workforce of cardiologists plus cardiology NPs and PAs. Columns follow Table S15; the final column notes overlap with the primary top 25 (Table S2).

| Rank | County | State | Tract FIPS | Cardiology NP/PA in county | DDI composite | Burden z score | In primary top 25 |
| --- | --- | --- | --- | --- | --- | --- | --- |
| 1 | Marion | IN | 18097357602 | 103 | 15.00 | 1.45 | Yes |
| 2 | Chatham | GA | 13051003302 | 8 | 16.00 | 1.46 | Yes |
| 3 | Henrico | VA | 51087201405 | 19 | 17.30 | 1.49 | Yes |
| 4 | Mobile | AL | 01097005100 | 28 | 13.50 | 1.14 | Yes |
| 5 | Milwaukee | WI | 55079004000 | 25 | 17.10 | 1.45 | Yes |
| 6 | Marion | FL | 12083001206 | 29 | 18.40 | 1.53 | Yes |
| 7 | Miami-Dade | FL | 12086014100 | 87 | 17.70 | 1.42 | Yes |
| 8 | Harris | TX | 48201331603 | 137 | 18.50 | 1.48 | Yes |
| 9 | Clayton | GA | 13063040536 | 2 | 11.60 | 0.87 | Yes |
| 10 | Tunica | MS | 28143950102 | 0 | 18.30 | 1.45 | Yes |
| 11 | Wayne | MS | 28153950301 | 0 | 14.80 | 1.11 | Yes |
| 12 | Marion | IN | 18097381207 | 103 | 11.80 | 0.84 | Yes |
| 13 | Marion | IN | 18097340600 | 103 | 17.90 | 1.38 | Yes |
| 14 | Harris | TX | 48201453702 | 137 | 15.10 | 1.08 | Yes |
| 15 | St. Louis | MO | 29189210501 | 102 | 18.00 | 1.32 | Yes |
| 16 | Prince George's | MD | 24033803001 | 19 | 14.90 | 1.02 | Yes |
| 17 | Marion | IN | 18097310306 | 103 | 16.30 | 1.14 | Yes |
| 18 | Cabarrus | NC | 37025040705 | 77 | 7.50 | 0.35 | Yes |
| 19 | Harris | TX | 48201550305 | 137 | 8.20 | 0.41 | Yes |
| 20 | Lee | AL | 01081041702 | 4 | 17.00 | 1.19 | Yes |
| 21 | Honolulu | HI | 15003009803 | 15 | 16.40 | 1.14 | Yes |
| 22 | Blount | TN | 47009010100 | 14 | 16.10 | 1.10 | Yes |
| 23 | Hernando | FL | 12053041602 | 11 | 18.70 | 1.33 | Yes |
| 24 | Broward | FL | 12011060302 | 48 | 16.60 | 1.15 | Yes |
| 25 | Mobile | AL | 01097006903 | 28 | 15.40 | 1.01 | Yes |

**Table S17.**
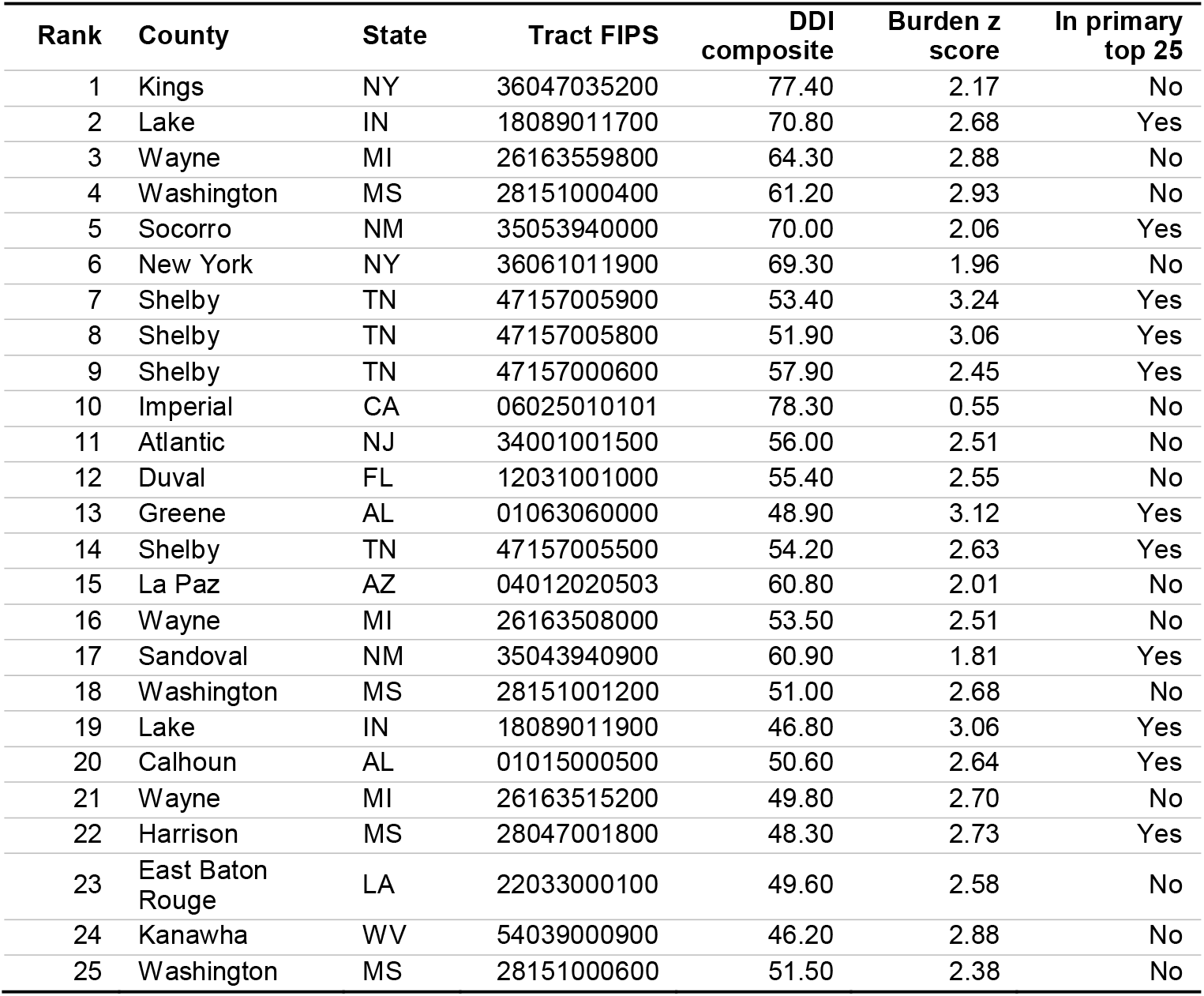
Top 25 investment tracts with the workforce-constrained pool rebuilt from the primary care physician workforce, counties with zero primary care physicians in 2023 or a declining per-capita rate from 2020 to 2023. The final column flags tracts that also appear in the primary top 25 (Table S1).

**Table S18.**
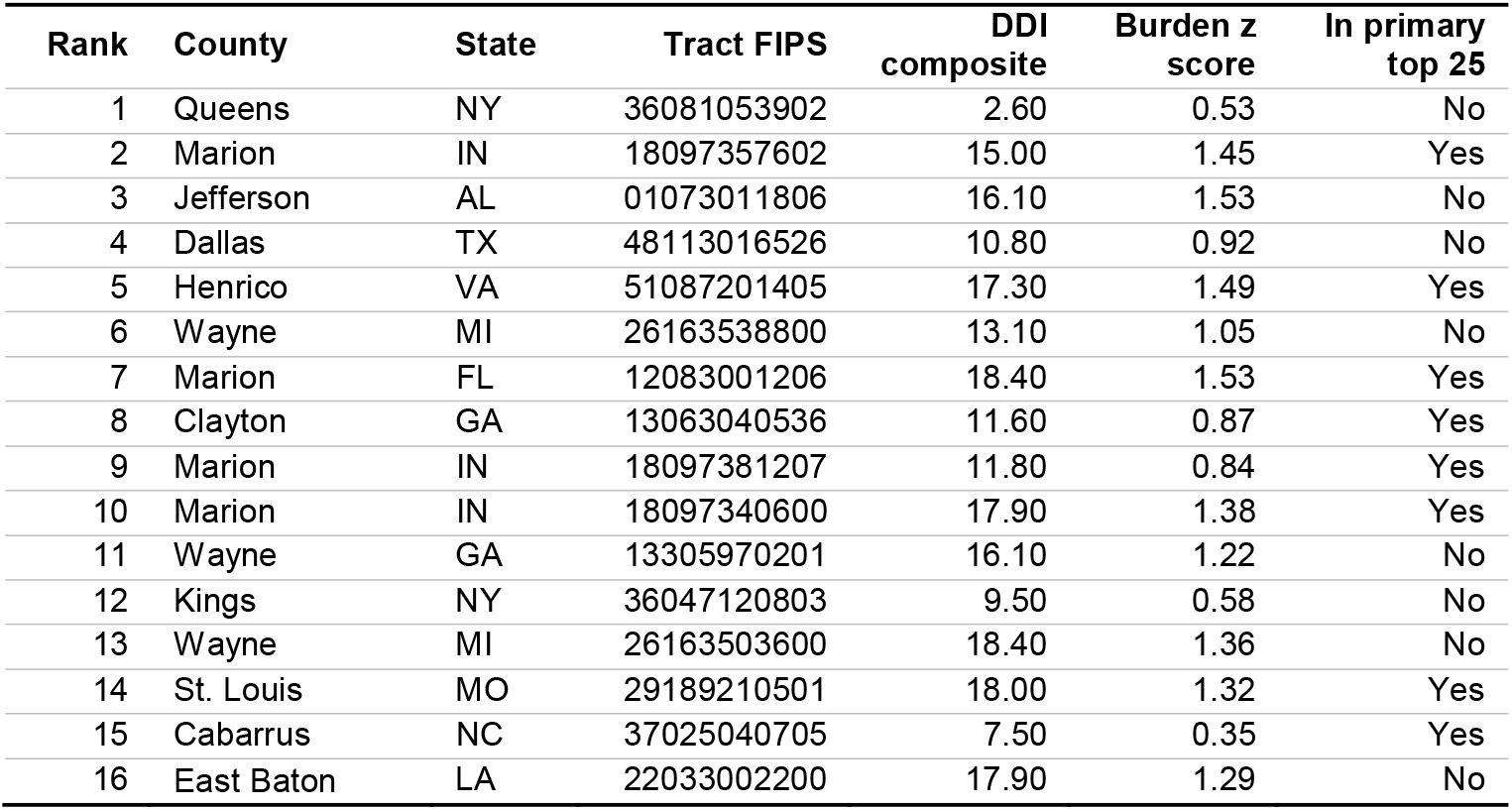
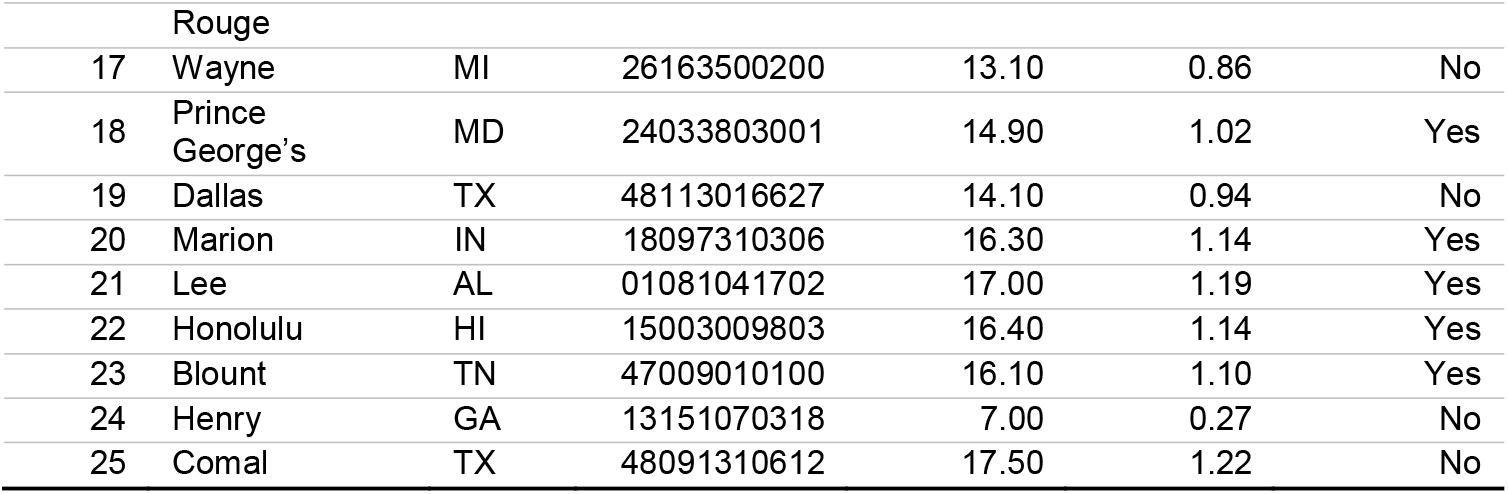
Top 25 deployment tracts with the workforce-constrained pool rebuilt from the primary care physician workforce. Columns follow Table S17, with the final column indicating overlap against the primary top 25 (Table S2).

### Data Missingness

Three gaps in the public datasets affected the analysis. First, the Area Health Resources Files carry no cardiology workforce counts for Connecticut, whose 9 planning regions replaced its counties in the federal geography in 2022; Connecticut therefore could not enter the workforce-constrained pool (**Fig. 1a**). Workforce counts were also unavailable for 28 Virginia independent cities, which the files combine with adjacent counties, and for Kalawao County, Hawaii. Second, PLACES suppressed most estimates for Kentucky and Pennsylvania because the 2 states did not collect enough Behavioral Risk Factor Surveillance System data to meet CDC inclusion requirements in the underlying survey years. Only 1 of the 10 burden measures (short sleep duration) was available for their 4,707 tracts, so burden estimates in these states rest on a single indicator. Third, the Divide Index was undefined for 523 of 83,522 tracts nationwide, typically tracts with little or no resident population; these tracts were excluded, leaving the 82,999 analyzed tracts.

